# Adverse Drug Reactions in Tuberculosis Treatment: Incidence, Duration and Resolution Pathways from a Mixed-Methods Patient-centric Study in India

**DOI:** 10.1101/2025.11.27.24319442

**Authors:** Ridhima Sodhi, Tunisha Kapoor, Vindhya Vatsyayan, Iti Seth, Harsh Chandra, Nishita Gill, Manoj Singh, Arnab Pal, Shamim Mannan

## Abstract

Adverse drug reactions (ADRs) remain a major barrier to successful tuberculosis (TB) treatment. They undermine adherence, prolong morbidity, and increase the risk of treatment failures and mortality. Yet, evidence on their incidence, duration, and management across diverse patient groups remains limited. We conducted a mixed-methods study to address this gap, using a representative sample of patients from six states in India. Specifically, we combined ethnographic observations and interviews with 40 patients and stakeholders across three districts with a quantitative survey of 2,000 randomly selected TB patients across eight districts. The ethnographic analysis revealed a novel taxonomy of ADRs, distinguishing *active ADRs* (acute, clinically urgent conditions), from *passive ADRs* (persistent, lower-intensity conditions) that quietly undermine adherence in later treatment phases. Passive ADRs such as skin darkening and fatigue typically warrant little clinical attention, yet their persistence makes them highly relevant for patient management strategies aimed at supporting adherence and achieving TB elimination. This finding was further contextualized and strengthened by quantitative analysis, which provided robust statistical insights into their incidence across diverse patient profiles. The quantitative analyses also reveal a near-universal burden of ADRs, with 86% of patients reporting at least one ADR (Mean = 3.1, SD 2.38). Women reported ADRs more frequently and for longer durations, particularly cutaneous ADRs, while elderly patients were more prone to gastrointestinal and musculoskeletal ADRs. Younger patients and women reported the highest prevalence of vomiting (41%), which emerged as the only independent predictor of unsuccessful treatment completion (OR = 0.39, 95% CI: 0.20–0.76). The overall number of ADRs was also strongly correlated with adverse treatment outcomes (OR = 0.88, 95% CI: 0.78–0.98). The active-passive taxonomy, along with risk-group profiling, offers a roadmap for differentiated counselling and pro-active patient-centric ADR management. We recommend embedding this approach into national TB protocols, with structured risk-based patient counselling at different stages of treatment, supported by adequate training for treatment coordinators and providers. While further research is warranted to assess scalability and cost-effectiveness, our findings demonstrate both the urgency and the feasibility of structured ADR management in high-burden TB settings.

**Author Summary:** Adverse drug reactions (ADRs) among drug-sensitive TB patients pose a significant challenge in TB treatment, particularly in resource-constrained settings. Yet, literature on their incidence, duration and resolutions pathways remains scarce - especially studies documenting these across socio-demographic profiles. Our study addresses this gap using a sequential mixed-methods design, combining qualitative and quantitative methodologies. The approach contributes two key advances to the literature. First, it introduces a novel taxonomy of ADRs, grounded in patients’ lived experiences and the support received. This taxonomy segregates ADRs into “active” ADRs, which are more severe and require urgent medical attention, and “passive” ADRs, which are less severe but persist for a longer time, and quietly undermine adherence. Second, it provides statistically validated evidence on the incidence, duration, and resolution of eleven ADRs, disaggregated across socio-demographic profiles. Together, these findings provide a roadmap for developing differentiated, risk-tailored protocols that emphasize gender- and age-responsive counselling. We recommend adapting these insights to inform dedicated ADR guidance within the National TB Elimination Frameworks (NTEP) for India, as well as other high-burden countries, clearly outlining the roles and responsibilities of healthcare providers - particularly treatment coordinators and physicians - and supporting them through targeted training led in collaboration with the NTEP.

## Introduction

Tuberculosis (TB) continues to be the leading cause of death due to infectious diseases worldwide, despite being preventable and curable [1,2]. More than 25% of the global TB incidence, and 28% of TB deaths, occur in India [2]. A range of initiatives towards TB elimination have been implemented over the years, across India and other high-burden countries [3–8]. These initiatives have included a varied set of approaches to drive adherence – from DOTS [9,10] to digital adherence technologies (DATs) [3–5,11]. However, adverse drug reactions (ADRs) of varying type and severity are known to affect adherence among those on anti-tubercular therapy (ATT) [12–15], even in patients on first line drugs (FLDs) [16–18]. Adherence gaps are a severe impediment to TB elimination goals – as they prolong treatment, contribute to drug-resistance, increase loss to follow up, and ultimately increase TB incidence and mortality [19–21]. Even as majority of TB patients are on FLDs, evidence continues to focus on drug-resistant TB patients, or those on second-line drugs (SLDs) [18,20,22–24]. Studies quantifying incidence and duration of ADRs among drug-sensitive TB (DS-TB) patients are particularly scarce [18,25] and rarely report across diverse socio-demographic groups, or document patient-driven resolution strategies.

Understanding ADR patterns among this group is critical, but challenging, especially in high-burden countries like India, Indonesia, the Philippines, Pakistan, and Nigeria, where under-reporting, limited patient support, and inadequate awareness persist [26–30]. Precise evidence on ADR patterns, however, can help support differentiated and cost-effective ADR management protocols, more suited in resource constrained settings.

Our study addresses this gap through a holistic assessment of patient experiences, leading to the identification of a novel active-passive taxonomy of ADRs, followed by an estimation of the precise incidence and duration of each one of those, combined with preferred resolution strategies. The insights, when looked to identify risk groups among patient profiles, hold important lessons on how to incorporate ADR management within NTEP protocols.

## Methods

### 2.1 Study design & Objective

We employed a mixed methods, non-experimental design, with an exploratory-sequential and theory-development approach [31,32] [Fig 1]. The first phase (April 2020 – June 2020) was ethnographic and examined patients’ lived experiences of TB. It included understanding the pre-diagnostic journey (e.g., finding the right doctor, diagnostic delays), stigma, treatment experience (ADRs and resolutions sought), and financial challenges. Including an observational and interview component, this phase informed a novel taxonomy of ADRs – active and passive ADRs. Insights from the qualitative phase informed the quantitative validation of ADR patterns across a statistically powered and representative sample of 2,000 TB patients (September 2020 - October 2020).

**Fig 1.**
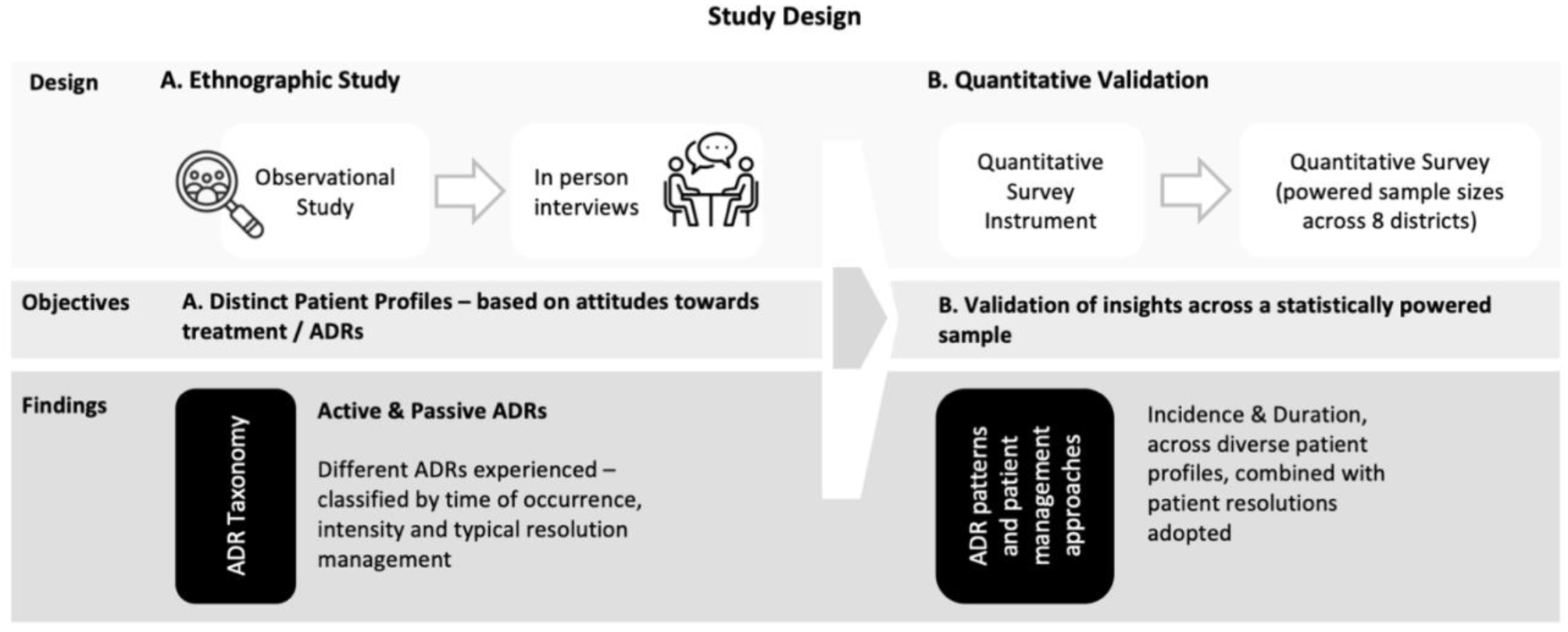
Methodological Overview.

### 2.2 Study Setting-Project JEET

The study was nested within Project JEET (Joint Effort for Elimination of TB), a large-scale private sector engagement initiative supported by the Global Fund and implemented in collaboration with the National TB Elimination Programme (NTEP) [33]. The qualitative phase of the study was conducted in three districts (Delhi, Bhopal and Jaipur), and the quantitative phase was conducted in additional five districts, totaling eight districts in total (Delhi, Gurgaon, Jaipur, Darbhanga, Indore, Bhopal, Ahmedabad, and Surat). The sites were so chosen as the William J. Clinton Foundation was managing the Patient Provider Support Agency (PPSA) under the JEET program [33,34].

### 2.3 Stakeholders

Six stakeholder groups were included: a) patients, b) HCPs, c) hub agents, d) treatment coordinators (TCs), e) field staff, and f) project / program directors. In the PPSA model, hub agents facilitated timely diagnosis, referrals, sample collection, transport, and patient assignment to TCs [33]. TCs were responsible for monthly follow-ups, counselling, ADR guidance, psychosocial support, and ensuring prescription refills when government-supplied FDCs were used [33]. While the qualitative phase included all six stakeholder groups, the quantitative phase focused on patients and TCs.

### 2.4 Sample selection

**Qualitative** sampling relied on a purposive approach – with two sub-components. The first component (<u>observational</u>) included observing five high-volume doctors (1 in Jaipur, 2 each in Delhi and Bhopal) in their routine clinical setting, alongside eight patients (2 in Jaipur, 2 in Delhi, and 4 in Bhopal) being treated in the same facilities, as well as their TCs and hub agents. The second component consisted of <u>in-person interviews</u> with 40 patients, 5 family members, 20 TCs, 12 HCPs, 8 hub agents, and 4 program staff across Delhi, Jaipur, and Bhopal [S1 Supp Text]. Patients were purposively selected to reflect socio-demographic diversity (age, gender, marital status, occupation) and treatment stage: (a) intensive phase (<2 months), (b) continuation phase (≥2 months completed), and (c) post-treatment completion. For the **quantitative** phase, a random selection method without replacement, combined with proportionate stratified sampling, was used to achieve representation across age, gender, site of disease, and treatment stage. Sample sizes for each geography were calculated at 95% confidence with a 5% margin of error, assuming a 50% response distribution, using the ‘*rsampcalc*’ function of the ‘*sampler’* package in R Studio.

### 2.5 Data Collection

Qualitative data were collected through (a) observations in clinics and hospitals, with field notes on communication, behaviors, and latent influences, and (b) semi-structured interviews. One researcher led the discussion; another documented through notes, photographs, and sketches. In the quantitative phase, structured patient interviews were administered by designated TCs, leveraging their rapport with patients. TCs underwent a two-day training, administered in both Hindi and English, covering objectives, ethics, and standardized protocols, followed by a pilot. Due to COVID-19 restrictions, surveys were conducted telephonically. To minimize bias, questions focused on treatment journeys and did not probe the quality of TC counselling. TCs were also surveyed digitally, though those findings are not assessed for this study.

### 2.6 Survey Instruments

The qualitative phase used semi-structured discussion guides [S2 Supp Text], informed by literature, discussions with program staff, and observational insights. For patients, guides explored sociodemographic, pre-diagnosis pathways, TB knowledge and perceptions, treatment regimens, ADR experiences, adherence behaviours, support from TCs and family, and the broader impact of TB on daily life. Questions elicited both factual (e.g., pill counts, treatment duration) and experiential (e.g., ADR coping, missed doses, stigma) responses, ensuring coverage of both clinically recognised ADRs and patient-driven perspectives.

Themes from qualitative interviews informed the quantitative instrument. Modules covered demographics, pre-diagnosis journey, treatment-related challenges, and attitudes towards treatment. ADRs were explicitly categorized as active or passive, with separate closed-ended questions on incidence and duration of individual ADRs, and a general question on how ADRs were managed. Patients could also report additional ADRs or management strategies in open-text fields, later re-coded. This ensured systematic capturing of both structured data and patient narratives.

### 2.7 Data Sources & Eligibility

The primary data source were in-person (qualitative) and telephonic interviews (quantitative). The secondary source was programmatic data collected by JEET staff, collected as part of services rendered under the project, and used for sampling calculations. This was used to supplement information on socio-demographic (age, sex, district) and treatment related (diagnostic test, type of diagnosis - pulmonary or extrapulmonary, free drug provision, and treatment outcomes) covariates. Definitions of outcomes are detailed in S1 Appendix, S1 Table.

### 2.7 Inclusions and exclusions

The study was conducted across eight JEET districts (Delhi, Gurgaon, Jaipur, Bhopal, Indore, Darbhanga, Ahmedabad, and Surat), with qualitative interviews and observations limited to Delhi, Jaipur, and Bhopal. The study population comprised adult (≥16 years) pulmonary drug-sensitive TB patients, as pediatric patients (≤15 years) were excluded to account for differences in treatment regimens. Only patients assigned a treatment coordinator (TC) were included, and those with drug-resistant TB were excluded due to differing and more complex regimens. To minimize recall bias, patients with more than 22 weeks of ongoing treatment were excluded from the quantitative survey, as their ability to accurately recall ADRs is expected to be lower. In contrast, the qualitative phase purposively included patients across all three treatment stages - intensive, continuation, and post-treatment completion - to capture a broader range of experiences.

**Fig 2:**
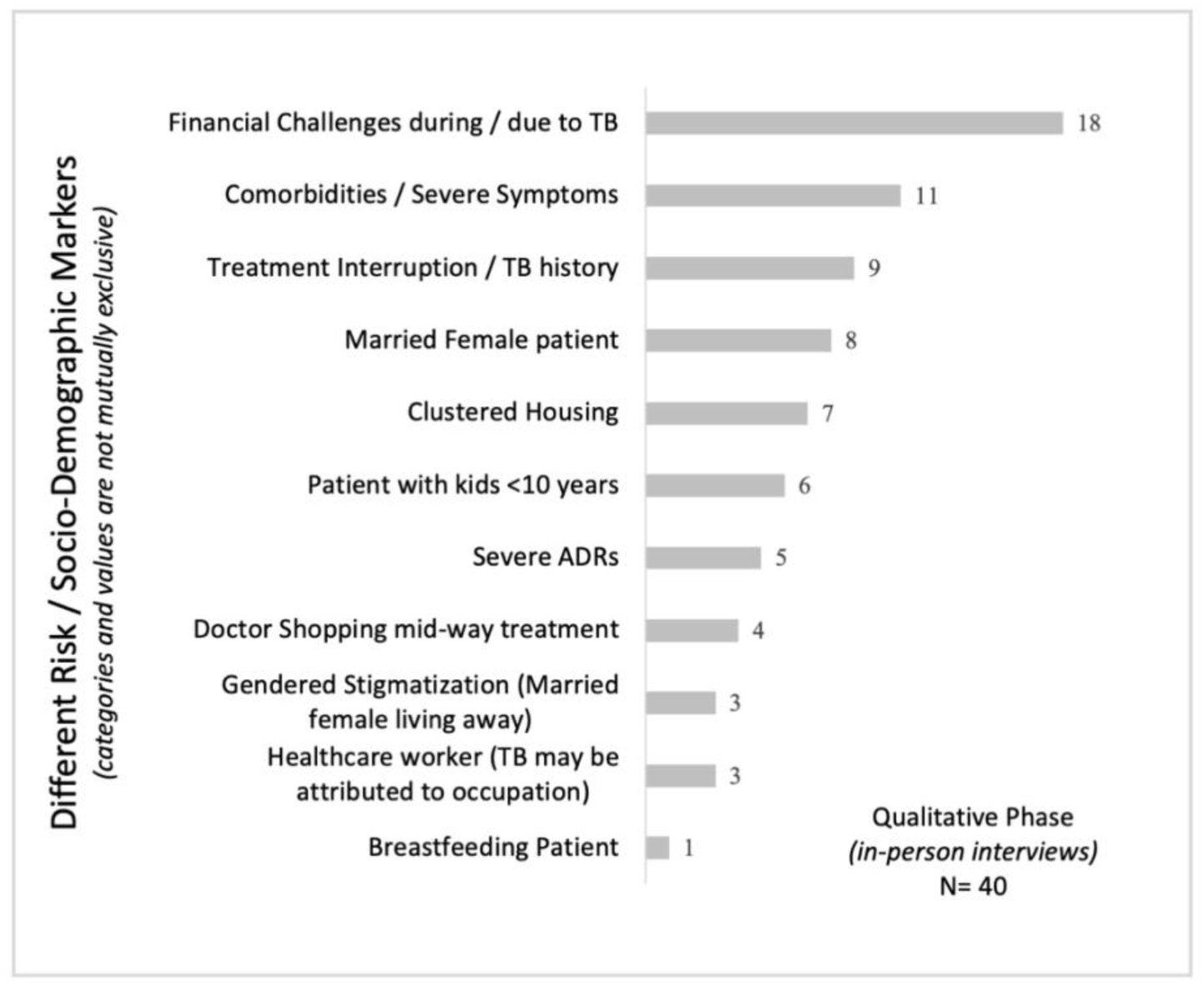
Distribution of patient profiles identified during the qualitative phase. Note: Categories are thematically grouped and not mutually exclusive.

### 2.8 Data Consolidation and Confidentiality

Multiple steps were taken to ensure data safety and participant confidentiality. Audio recordings were stored in a password-protected hard drive, accessible only to the primary research staff, and transcripts were de-identified prior to the analysis. For the quantitative phase, surveyed patients were assigned a new identification ids; the matching identification for linkage with programmatic data (treatment outcomes) was maintained separately by program staff. The merged data was re-encrypted using a UUID (universally unique identifier) before being used for research.

### 2.6 Ethics Approval

Institutional ethics clearance was obtained from the Centre for Media Studies Institutional Review Board (CMS-IRB), New Delhi (Reference ID: IRB00006230). The recruitment period for the study was between 13^th^ April 2020 and 15^th^ October 2020. Informed consent was obtained in writing for in-person interviews and verbally for telephonic surveys, with verbal consent digitally recorded. No minors were included.

### 2.7 Software & Analysis

#### Qualitative

Interviews were transcribed into dialogue-style transcripts in Trello, cross-checked for accuracy, and coded by theme (behaviors, attitudes, actions). Field notes were tagged similarly for consistency.

#### Quantitative

Analysis was conducted in R Studio (2022.07.02). Sample sizes were determined using rsampcalc. Descriptive statistics were complemented with Mann–Whitney U, Kruskal–Wallis, and Sidak tests for group differences, and logistic regression models for associations. p < 0.05 was considered statistically significant.

## Results

### Qualitative findings

A total of 40 patients, with a fairly equal gender-split (men=19, women=21), were interviewed in the first phase. Primary findings from this phase, along with recommended ADR management strategies are illustrated in Fig 3.

**Fig 3:**
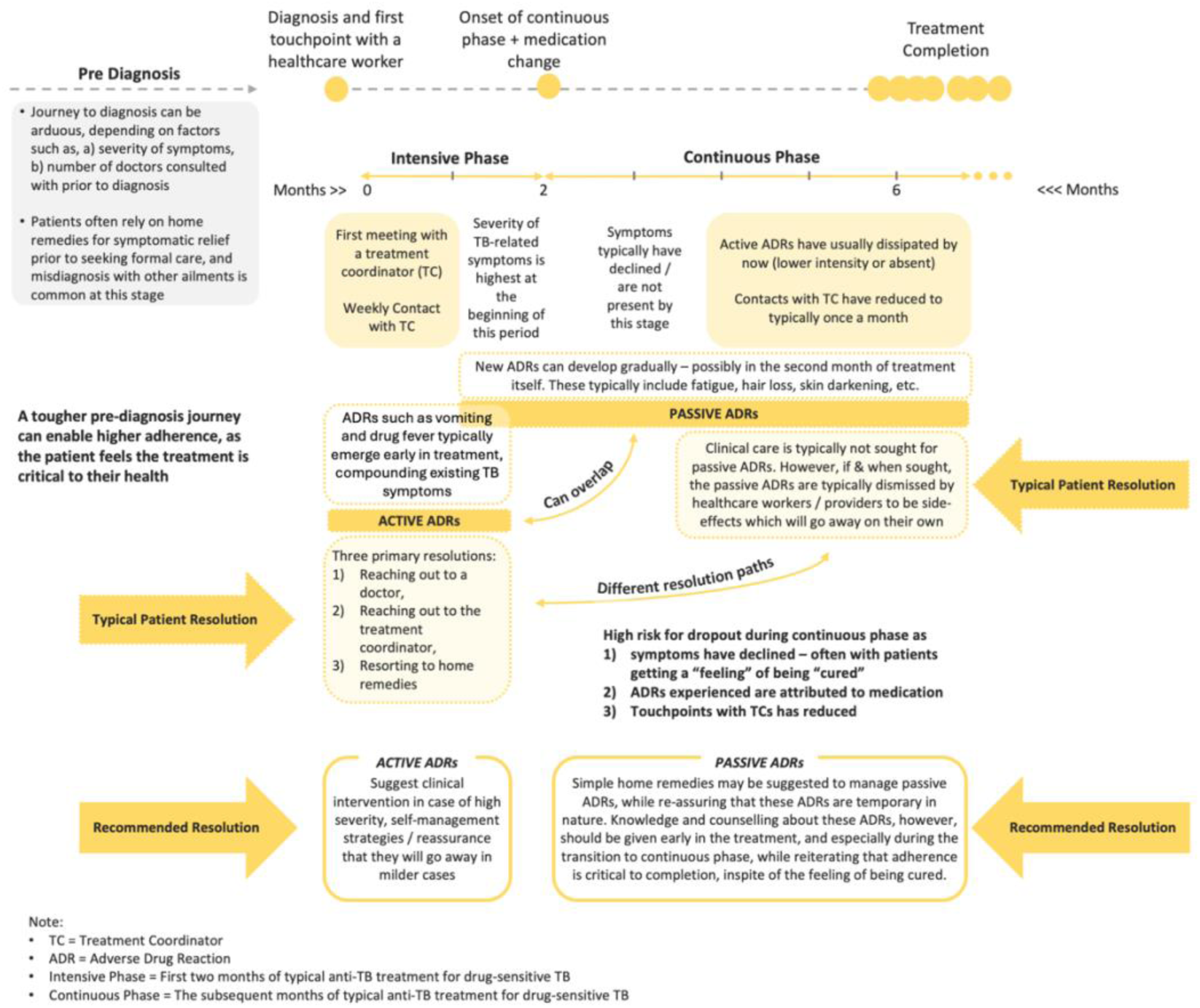
Patient Pathway, ADRs and Recommended management strategies.

#### Pre-Diagnosis Journey

Typical symptoms experienced by patients prior to seeking a resolution include cough, persistent fever, and chest pain. Initial reliance on home remedies is common, as is misdiagnoses by HCPs, which prolongs the diagnostic pathway. Typical challenges include delay or/and difficulty in accessing a doctor, as well as reliance on clinical diagnosis instead of advanced diagnostics due to availability or/and inability to pay. Prolonged endurance of symptoms exacerbates the pre-diagnosis journey – which is correlated with patients adopting a more serious attitude towards their treatment, often leading to better adherence.

#### ADR taxonomy – Active and Passive ADRs

While the onset of TB treatment corresponds to a decline in TB-related symptoms, it also coincides with ADRs for most patients. While many ADRs subside within the first few weeks of treatment, others appear later and may persist for extended periods. These late-occurring ADRs can confuse patients, who may misinterpret them as signs of recovery and justification to discontinue or skip medications.

Traditional ADR classifications, grounded in biomedical definitions, focus on clinical characteristics such as type (gastrointestinal, dermatological), and severity (mild, moderate, severe). Building on patient experiences, our study introduces a novel taxonomy categorizing ADRs as active or passive using three patient-centric dimensions, 1) typical time of occurrence, 2) intensity, and 3) resolutions sought.

##### Active ADRs

These are ADRs (drug fever, joint pains, diarrhoea, itching, vomiting, and trepidation) most commonly known and experienced, typically exhibiting in the initial stages of treatment. Typically dealt with an “active” intervention, owing to their relative intense experience, these are termed as “Active ADRs”. While they warrant support from a medical professional, patients also reach out to TCs for assistance. Due to the timing and intensity, these ADRs can lead the patient to believe the diagnosis to be incorrect, posing a risk to treatment continuation.

##### Passive ADRs

TB-related symptoms, as well as active ADRs typically subside by the end of the intensive period, leaving some patients with the perception of being “treated”. A risk factor on its own, this perception is sometimes complemented by a gradual emergence of certain other ADRs like fatigue, hair fall, acne, skin darkening, and teeth darkening. Typically mild, patients often do not seek an immediate redressal for these ADRs – which tend to persist over longer durations. If consulted, HCPs typically ask the patients to wait for these ADRs to subside, or endure them until the end of the treatment. Alternative practitioners are often sought, apart from home remedies. Classified as passive ADRs, these pose a significant risk to treatment completion as the patient is likely to pay less importance to medication at this stage, unless adequately counselled. These ADRs were not found in the NTEP (National TB Elimination Program) documentation [35].

> Original Quote (Hindi): “Yeh khaali pet waali davai se jee machalta tha, mann karta tha nahi le” Translated: “Taking this medicine on an empty stomach made me feel nauseous; I felt like not taking it.”

#### Resolution pathway for ADR Management

Three resolution pathways are observed, 1) home remedies, 2) reaching out to the TC, and 3) consulting with treating HCP or alternative HCP. Of these, home remedies are not always effective. The other alternative is to reach out to TCs, whose training did not allow them to recommend or prescribe treatments. Hence, they typically advised patients to seek advice from their treating HCPs. The HCPs, in turn, often recommended waiting for the ADRs to subside on their own, unless the symptoms are considered particularly severe. In the latter case, which is less common, alternative measures are pursued, such as modifying the medication regimen, referring the patient to a specialist, or initiating additional clinical investigations.

> Original Quote (Hindi): “doctor ne pehle bataya tha ki aisa hoga, aur phir khud theek ho jaayega” Translated: “The doctor had told me beforehand that this would happen, and that it would get better on its own.”

#### Identified Patient profiles

Based on differences in attitudes towards treatment, ADR management, and identified risk factors, three patient profiles were identified in the qualitative phase (Fig 4). <u>Caregivers</u>, largely women supporting both younger and elderly family members, confirmed lack of available support to them, in addition to additional stress around how the diagnosis may affect their caregiving responsibilities. <u>Wage earners</u> (primary or secondary) were specifically concerned with the financial consequences of the diagnosis and its impact on household stability. <u>Dependents</u> had fewer responsibilities and were primarily focused on their own health and recovery. Across these groups, even seemingly mild ADRs threatened adherence, particularly for those balancing financial responsibilities or caregiving roles, where ADRs directly interfered with their ability to work or provide care.

**Fig 4:**
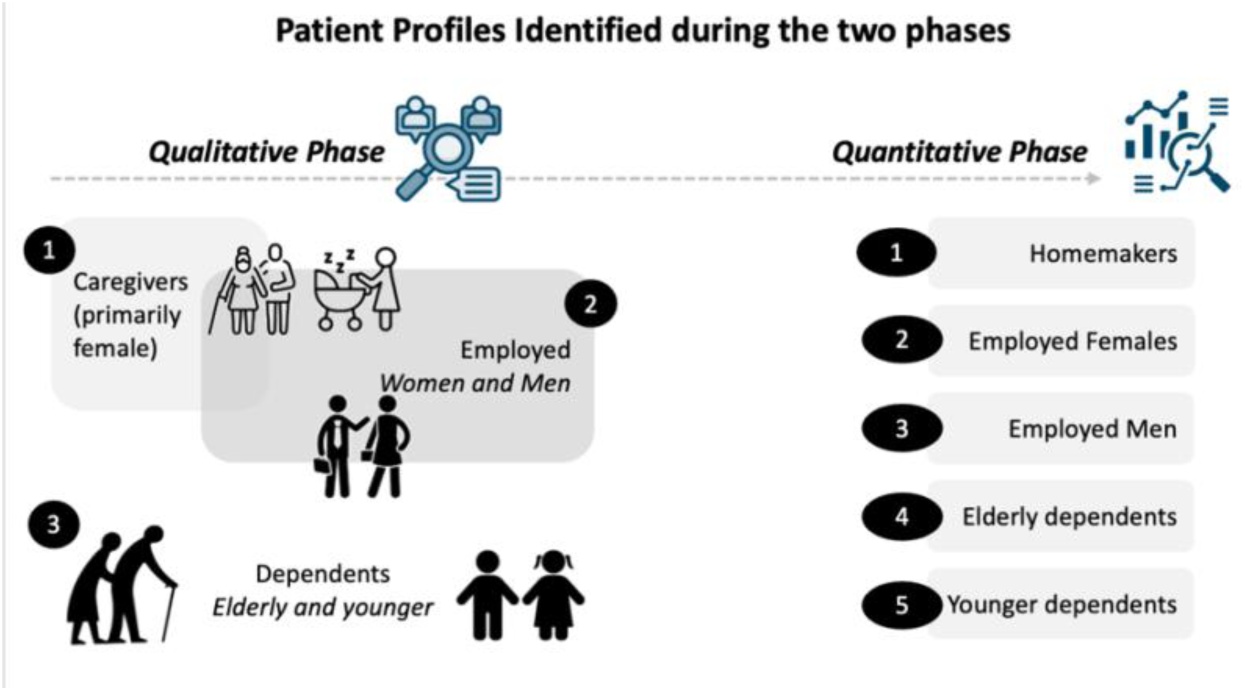
Classification of TB patients, developed from qualitative findings and validated through quantitative data.

These categories were revised to five for quantitative validation, considering the differential risk factors based on gender and age (Fig 4).

### Quantitative findings

Table 1 summarizes patient characteristics by age, gender, diagnosing district, and TB type (pulmonary or extra pulmonary). Among 2,124 survey responses, 55% were males, and 31% had extra pulmonary TB. Eleven different ADRs were assessed, and an individual was found to typically experience 3 ADRs [Mean = 3.11, Median = 3, IQR: 1,5]. Five different patient categories were reported for, drawn from patient profiles identified in the qualitative study [Table 2, Fig 4]. Summary statistics for the five profiles are reported in S2 Table (S1 Appendix).

**Table 1:**
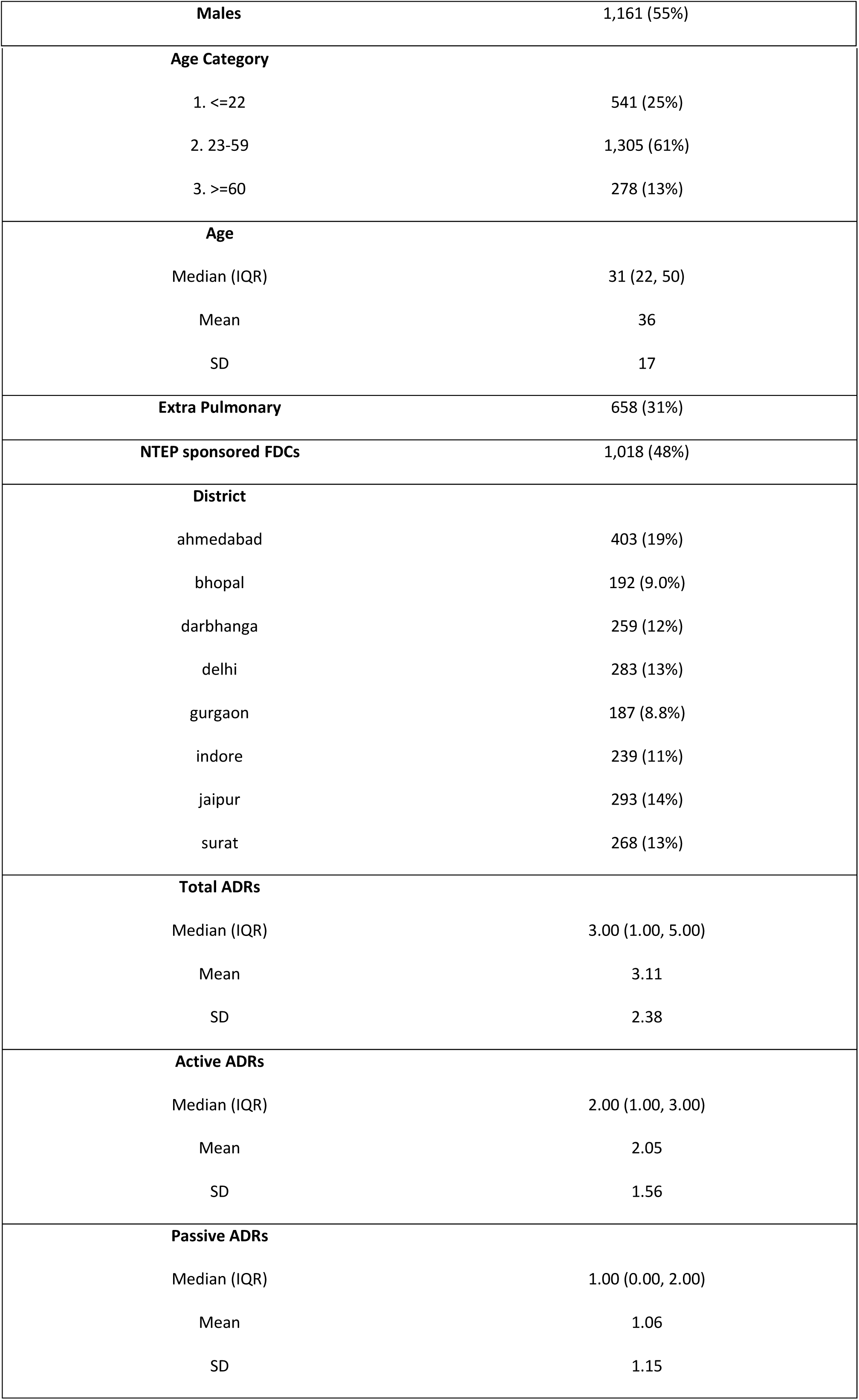
Summary Statistics for patients surveyed; N = 2,124.

**Table 2:**
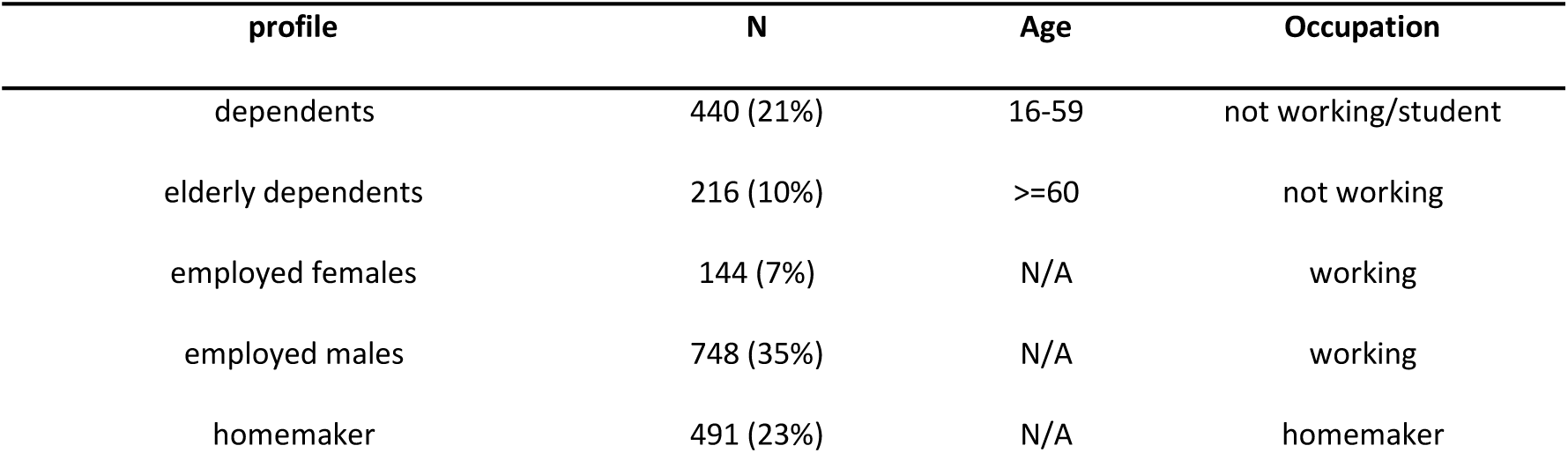
Categorization of patients by gender, age, and occupational status; N = 2,039.

| profile | N | Age | Occupation |
| --- | --- | --- | --- |
| dependents | 440 (21%) | 16-59 | not working/student |
| elderly dependents | 216 (10%) | >=60 | not working |
| employed females | 144 (7%) | N/A | working |
| employed males | 748 (35%) | N/A | working |
| homemaker | 491 (23%) | N/A | homemaker |

#### ADRs reported

Of the 11 ADRs assessed [Table 3], 86% of patients reported at least one, and 56% reported three or more [Table 4]. The most common ADRs were fatigue (55%) and trepidation (51%), followed by joint pain (46%), drug fever (40%), and vomiting (34%). Skin (17%), hair (20%), and dental challenges (6%) were reported by a significant, but smaller share of patients. Females are found to report a significantly higher number of ADRs, across all categories, except for diarrhoea and teeth darkening [Tables 3, 5]. Elderly patients (>=60 years of age) are significantly more likely to experience diarrhoea (15%) and joint pains (50%), and least likely to report acne (4%), hair loss (12%), or skin darkening (13%) [Table 6]. Mean differences by age group are detailed in S3 Table (S1 Appendix).

**Table 3:** Individual ADRs experienced by patients, segregated by gender; N = 2,124.

| Type | ADR | Total Patients<br>N = 2124 | Females<br>N = 963 | Males<br>N = 1161 |
| --- | --- | --- | --- | --- |
| active | diarrhoea | 8% (180) | 8% (79) | 9% (101) |
| active | drug fever | 40% (839) | 42% (406) | 37% (433) |
| active | itching | 25% (541) | 29% (275) | 23% (266) |
| active | joint pains | 46% (983) | 50% (478) | 43% (505) |
| active | trepidation | 51% (1076) | 59% (565) | 44% (511) |
| active | vomit | 34% (730) | 41% (397) | 29% (333) |
| passive | acne | 9% (187) | 11% (107) | 7% (80) |
| passive | fatigue | 55% (1171) | 59% (570) | 52% (601) |
| passive | hair loss | 20% (417) | 30% (292) | 11% (125) |
| passive | skin darkening | 17% (353) | 19% (184) | 15% (169) |
| passive | teeth darkening | 6% (126) | 6% (59) | 6% (67) |

**Table 4:** Total number of ADRs reported; segregated by gender; N = 2,124.

| ADRs | Total Patients<br>N = 2124 | Females<br>N = 963 | Males<br>N = 1161 |
| --- | --- | --- | --- |
| 0 | 297 (14%) | 103 (11%) | 194 (17%) |
| 1 | 316 (15%) | 120 (12%) | 196 (17%) |
| 2 | 337 (16%) | 130 (13%) | 207 (18%) |
| 3 | 352 (17%) | 163 (17%) | 189 (16%) |
| 4 | 289 (14%) | 140 (15%) | 149 (13%) |
| 5 | 218 (10%) | 115 (12%) | 103 (9%) |
| 6 | 117 (6%) | 63 (7%) | 54 (5%) |
| 7 | 96 (5%) | 66 (7%) | 30 (3%) |
| 8 | 38 (2%) | 29 (3%) | 9 (1%) |
| 9 | 25 (1%) | 14 (1%) | 11 (1%) |
| 10 | 21 (1%) | 10 (1%) | 11 (1%) |
| 11 | 18 (1%) | 10 (1%) | 8 (1%) |

**Table 5:**
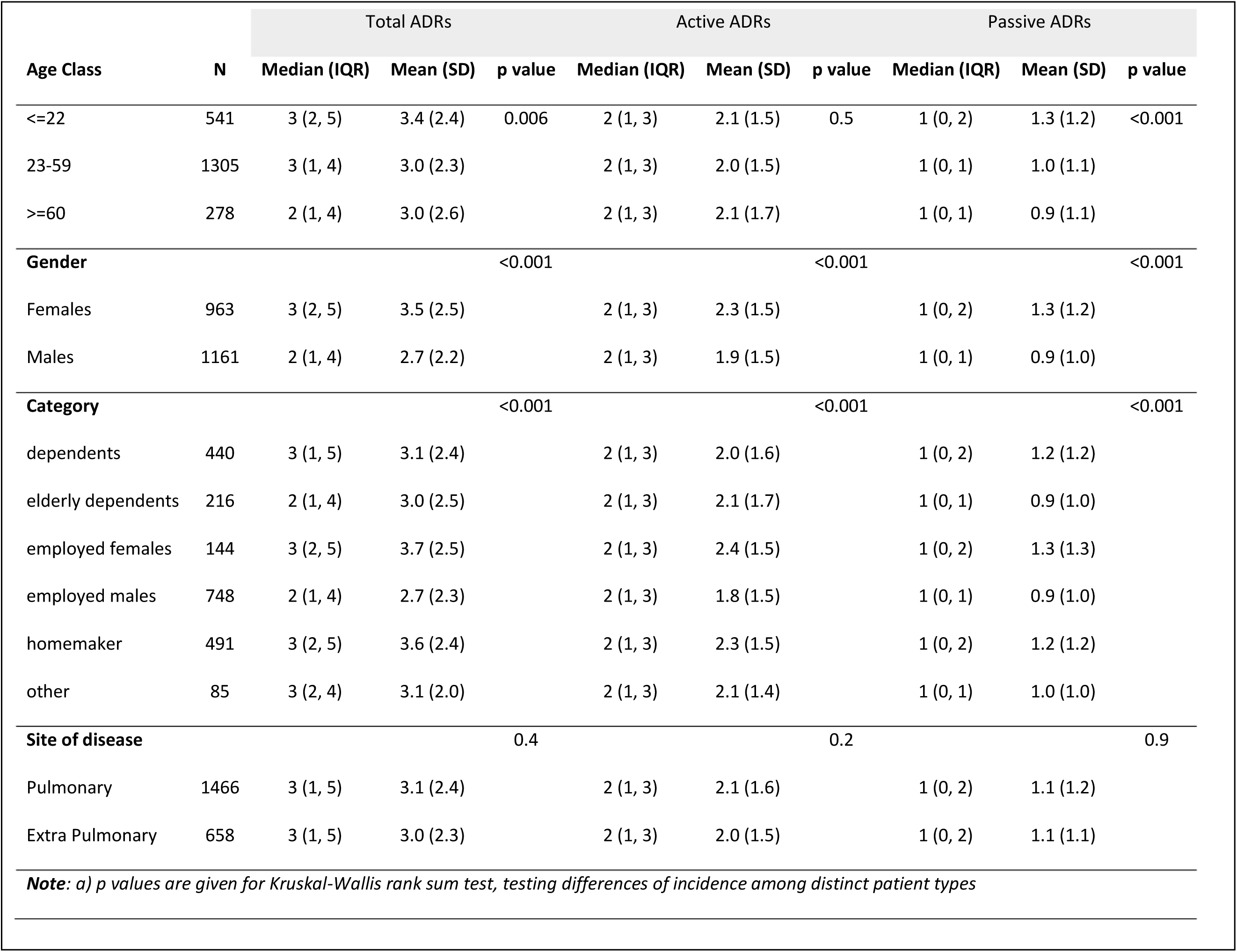
Summary statistics for number of ADRs reported; N = 2,124.

| Age Class | N | Total ADRs |  |  |  | Active ADRs |  |  |  | Passive ADRs |  |  |
| --- | --- | --- | --- | --- | --- | --- | --- | --- | --- | --- | --- | --- |
|  |  | Median (IQR) | Mean (SD) | p value |  | Median (IQR) | Mean (SD) | p value |  | Median (IQR) | Mean (SD) | p value |
| <=22 | 541 | 3 (2, 5) | 3.4 (2.4) | 0.006 |  | 2 (1, 3) | 2.1 (1.5) | 0.5 |  | 1 (0, 2) | 1.3 (1.2) | <0.001 |
| 23-59 | 1305 | 3 (1, 4) | 3.0 (2.3) |  |  | 2 (1, 3) | 2.0 (1.5) |  |  | 1 (0, 1) | 1.0 (1.1) |  |
| >=60 | 278 | 2 (1, 4) | 3.0 (2.6) |  |  | 2 (1, 3) | 2.1 (1.7) |  |  | 1 (0, 1) | 0.9 (1.1) |  |
| **Gender** |  |  |  | <0.001 |  |  |  | <0.001 |  |  |  | <0.001 |
| Females | 963 | 3 (2, 5) | 3.5 (2.5) |  |  | 2 (1, 3) | 2.3 (1.5) |  |  | 1 (0, 2) | 1.3 (1.2) |  |
| Males | 1161 | 2 (1, 4) | 2.7 (2.2) |  |  | 2 (1, 3) | 1.9 (1.5) |  |  | 1 (0, 1) | 0.9 (1.0) |  |
| **Category** |  |  |  | <0.001 |  |  |  | <0.001 |  |  |  | <0.001 |
| dependents | 440 | 3 (1, 5) | 3.1 (2.4) |  |  | 2 (1, 3) | 2.0 (1.6) |  |  | 1 (0, 2) | 1.2 (1.2) |  |
| elderly dependents | 216 | 2 (1, 4) | 3.0 (2.5) |  |  | 2 (1, 3) | 2.1 (1.7) |  |  | 1 (0, 1) | 0.9 (1.0) |  |
| employed females | 144 | 3 (2, 5) | 3.7 (2.5) |  |  | 2 (1, 3) | 2.4 (1.5) |  |  | 1 (0, 2) | 1.3 (1.3) |  |
| employed males | 748 | 2 (1, 4) | 2.7 (2.3) |  |  | 2 (1, 3) | 1.8 (1.5) |  |  | 1 (0, 1) | 0.9 (1.0) |  |
| homemaker | 491 | 3 (2, 5) | 3.6 (2.4) |  |  | 2 (1, 3) | 2.3 (1.5) |  |  | 1 (0, 2) | 1.2 (1.2) |  |
| other | 85 | 3 (2, 4) | 3.1 (2.0) |  |  | 2 (1, 3) | 2.1 (1.4) |  |  | 1 (0, 1) | 1.0 (1.0) |  |
| **Site of disease** |  |  |  | 0.4 |  |  |  | 0.2 |  |  |  | 0.9 |
| Pulmonary | 1466 | 3 (1, 5) | 3.1 (2.4) |  |  | 2 (1, 3) | 2.1 (1.6) |  |  | 1 (0, 2) | 1.1 (1.2) |  |
| Extra Pulmonary | 658 | 3 (1, 5) | 3.0 (2.3) |  |  | 2 (1, 3) | 2.0 (1.5) |  |  | 1 (0, 2) | 1.1 (1.1) |  |
| **Note:** a) p values are given for Kruskal-Wallis rank sum test, testing differences of incidence among distinct patient types |  |  |  |  |  |  |  |  |  |  |  |  |

**Table 6:** Summary statistics for Active ADRs reported; N = 2,124.

| Age Class | N | diarrhoea | p-value | drug fever | p-value | itching | p-value | joint pains | p-value | trepidation | p-value | vomit | p-value |
| --- | --- | --- | --- | --- | --- | --- | --- | --- | --- | --- | --- | --- | --- |
| <=22 | 541 | 7% | <0.001 | 44% | 0.032 | 29% | 0.09 | 39% | <0.001 | 49% | 0.6 | 41% | <0.001 |
| 23-59 | 1305 | 8% |  | 38% |  | 24% |  | 48% |  | 51% |  | 32% |  |
| >=60 | 278 | 15% |  | 36% |  | 24% |  | 50% |  | 53% |  | 33% |  |
| **Gender** |  |  | 0.7 |  | 0.022 |  | 0.003 |  | 0.005 |  | <0.001 |  | <0.001 |
| Females | 963 | 8% |  | 42% |  | 29% |  | 50% |  | 59% |  | 41% |  |
| Males | 1161 | 9% |  | 37% |  | 23% |  | 43% |  | 44% |  | 29% |  |
| **Category** |  |  | 0.067 |  | 0.022 |  | 0.1 |  | <0.001 |  | <0.001 |  | <0.001 |
| dependents | 440 | 7% |  | 40% |  | 26% |  | 37% |  | 45% |  | 40% |  |
| elderly dependents | 216 | 14% |  | 35% |  | 24% |  | 50% |  | 53% |  | 35% |  |
| employed females | 144 | 8% |  | 51% |  | 31% |  | 50% |  | 56% |  | 40% |  |
| employed males | 748 | 9% |  | 37% |  | 23% |  | 46% |  | 44% |  | 25% |  |
| homemaker | 491 | 7% |  | 41% |  | 29% |  | 53% |  | 62% |  | 41% |  |
| other | 85 | 8% |  | 44% |  | 19% |  | 47% |  | 55% |  | 36% |  |
| **Site of disease** |  |  | 0.07 |  | 0.004 |  | 0.061 |  | 0.8 |  | >0.9 |  | 0.022 |
| Pulmonary | 1466 | 0.09 |  | 42% |  | 24% |  | 46% |  | 51% |  | 0.36 |  |
| Extra Pulmonary | 658 | 0.07 |  | 35% |  | 28% |  | 47% |  | 50% |  | 0.31 |  |
| **Note:** a) p values are given for Kruskal-Wallis rank sum test, testing differences of incidence among distinct patient types |  |  |  |  |  |  |  |  |  |  |  |  |  |

#### Differences between Active and Passive ADRs across patient groups

Significant differences in total, active, and passive ADRs were observed across age groups, gender, and patient categories, but not by type of TB [Table 5, Figs 5, 6]. Females and homemakers reported significantly more total, active, and passive ADRs than other patient groups. Younger people reported a higher number of passive ADRs, with a larger share reporting acne (14%), hair loss (28%) and skin-darkening (21%), relative to older age categories [Tables 5, 7].

**Fig 5:**
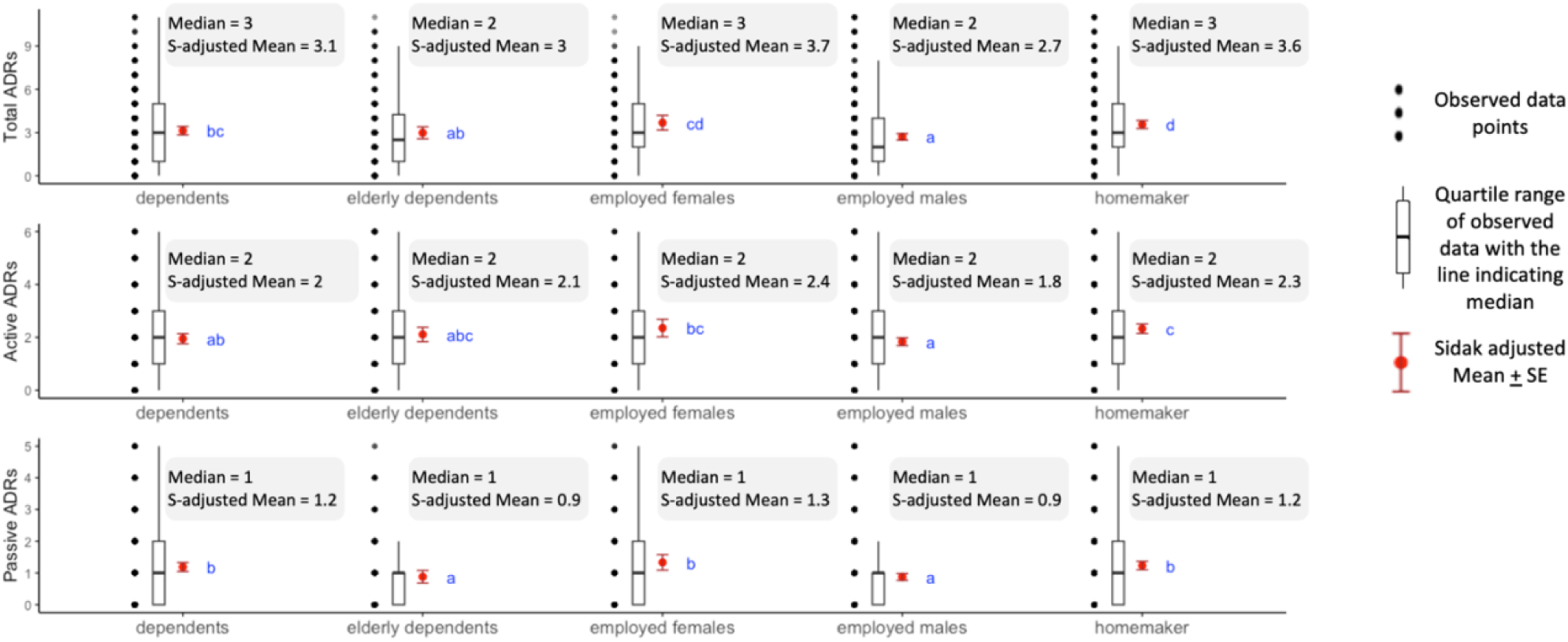
Box Plots for ADRs experienced by distinct profiles identified; N = 2,039. Notes: 1) Black dots represent raw data. Red dots and error bars represent (estimated marginal) means ± 95% confidence interval per group. Means not sharing any letter are significantly different by the Sidak-test at the 5% level of significance; 2) 85 patients with no work status defined are not categorized 2) Medians and Sidak-adjusted means are reported in figure text

**Fig 6:**
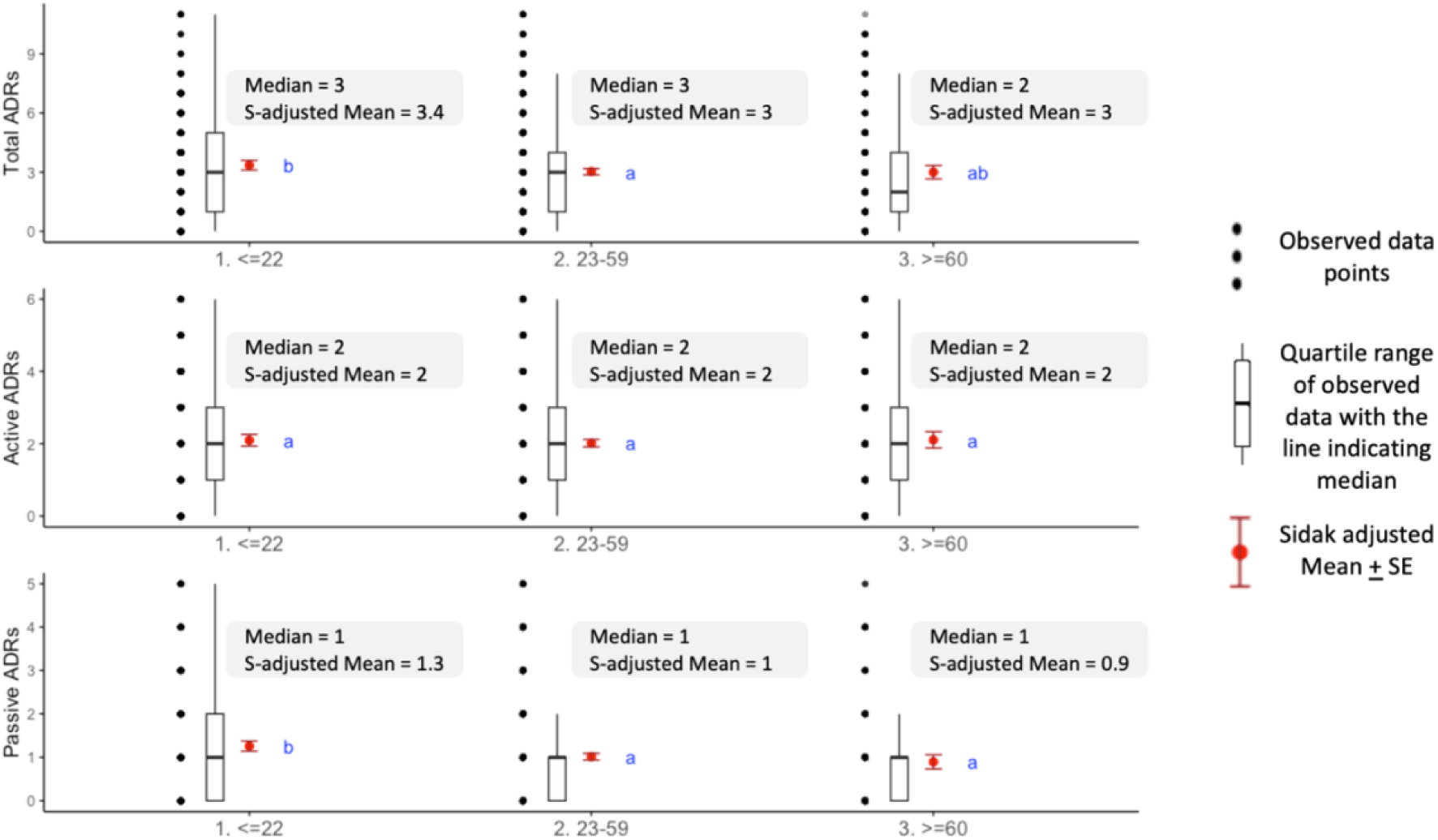
Box Plots for ADRs experienced by distinct age groups; N = 2,124. Notes: 1) Black dots represent raw data. Red dots and error bars represent (estimated marginal) means ± 95% confidence interval per group. Means not sharing any letter are significantly different by the Sidak-test at the 5% level of significance 2) Medians and Sidak-adjusted means are reported in figure text

**Table 7:** Summary statistics for Passive ADRs reported; N = 2,124.

| Age Class | N | acne | p-value | fatigue | p-value | hair loss | p-value | skin darkening | p-value | teeth darkening | p-value |
| --- | --- | --- | --- | --- | --- | --- | --- | --- | --- | --- | --- |
| <=22 | 541 | 14% | <0.001 | 57% | 0.3 | 28% | <0.001 | 21% | 0.002 | 5% | 0.034 |
| 23-59 | 1305 | 8% |  | 55% |  | 18% |  | 15% |  | 6% |  |
| >=60 | 278 | 4% |  | 51% |  | 12% |  | 13% |  | 9% |  |
| **Gender** |  |  | <0.001 |  | <0.001 | <0.001 | <0.001 |  | 0.005 |  | 0.7 |
| Females | 963 | 11% |  | 59% |  | 30% |  | 19% |  | 6% |  |
| Males | 1161 | 7% |  | 52% |  | 11% |  | 15% |  | 6% |  |
| **Category** |  |  | <0.001 |  | <0.001 |  | <0.001 |  | 0.081 |  | 0.8 |
| dependents | 440 | 14% |  | 54% |  | 27% |  | 19% |  | 6% |  |
| elderly dependents | 216 | 4% |  | 54% |  | 11% |  | 12% |  | 8% |  |
| employed females | 144 | 16% |  | 62% |  | 31% |  | 19% |  | 6% |  |
| employed males | 748 | 6% |  | 50% |  | 10% |  | 16% |  | 6% |  |
| homemaker | 491 | 9% |  | 61% |  | 30% |  | 19% |  | 6% |  |
| other | 85 | 9% |  | 66% |  | 14% |  | 12% |  | 4% |  |
| **Site of disease** |  |  | 0.6 |  | 0.11 |  | 0.5 |  | 0.3 |  | 0.3 |
| Pulmonary | 1466 | 0.09 |  | 56% |  | 19% |  | 16% |  | 6% |  |
| Extra Pulmonary | 658 | 0.09 |  | 53% |  | 21% |  | 18% |  | 5% |  |
| **Note:** a) p values are given for Kruskal-Wallis rank sum test, testing differences of incidence among distinct patient types |  |  |  |  |  |  |  |  |  |  |  |

#### Active ADRs; Duration

81% (1,729) patients reported experiencing some form of ADRs categorized as active, with only 4% to 22% experiencing any one of them for more than a month. While the length of experience varied among type of ADRs, most shared it to be up to 1 week [Table 8]. Drug fever and joint pains have the highest likelihood of being reported for a longer duration, with 41% and 45% patients reporting it to last for at least a month. Most patients who experience vomiting as an ADR report it to last for a week (84%), with 14% patients reporting it to last beyond a month. Patients diagnosed with extra pulmonary TB were found to experience diarrhoea for a longer duration with high statistical significance [S4 Table, S1 Appendix]. Women were found to typically report ADRs for slightly longer duration. Differences in duration were not found to be significant amongst other patient types.

**Table 8:** Duration of active ADRs; N = 1729.

| ADR | N | <=1 week | 2-3 weeks | 1 month | 1.5 months | 2 months | >=2 months |
| --- | --- | --- | --- | --- | --- | --- | --- |
| diarrhoea | 180 | 52% | 21% | 23% | 3% | 1% | 1% |
| drug fever | 839 | 34% | 25% | 19% | 4% | 6% | 12% |
| itching | 541 | 41% | 26% | 18% | 4% | 4% | 7% |
| joint pains | 983 | 31% | 24% | 23% | 5% | 7% | 10% |
| trepidation | 1076 | 47% | 26% | 16% | 4% | 3% | 4% |
| vomit | 730 | 64% | 22% | 8% | 2% | 2% | 2% |

**Note:** a) N represents the number of patients who experienced the ADR; b) subsequent values describe the duration of the ADR experienced by the illustrated share of patients

#### Passive ADRs; Duration

63% (1,333) patients reported experiencing some form of ADRs categorized as passive, with 18% to 52% experiencing any one of them for more than a month. Among these, hair loss was reported to last for the longest duration, with women significantly more likely to experience it for a longer time [Tables 9, 11]. For 70% of patients, fatigue was reported to last at least 1 month [Table 9]. Most (58%) patients who report teeth darkening shared it to last for less than 2 weeks. Site of disease was not found to be a significant differentiator.

**Table 9:**
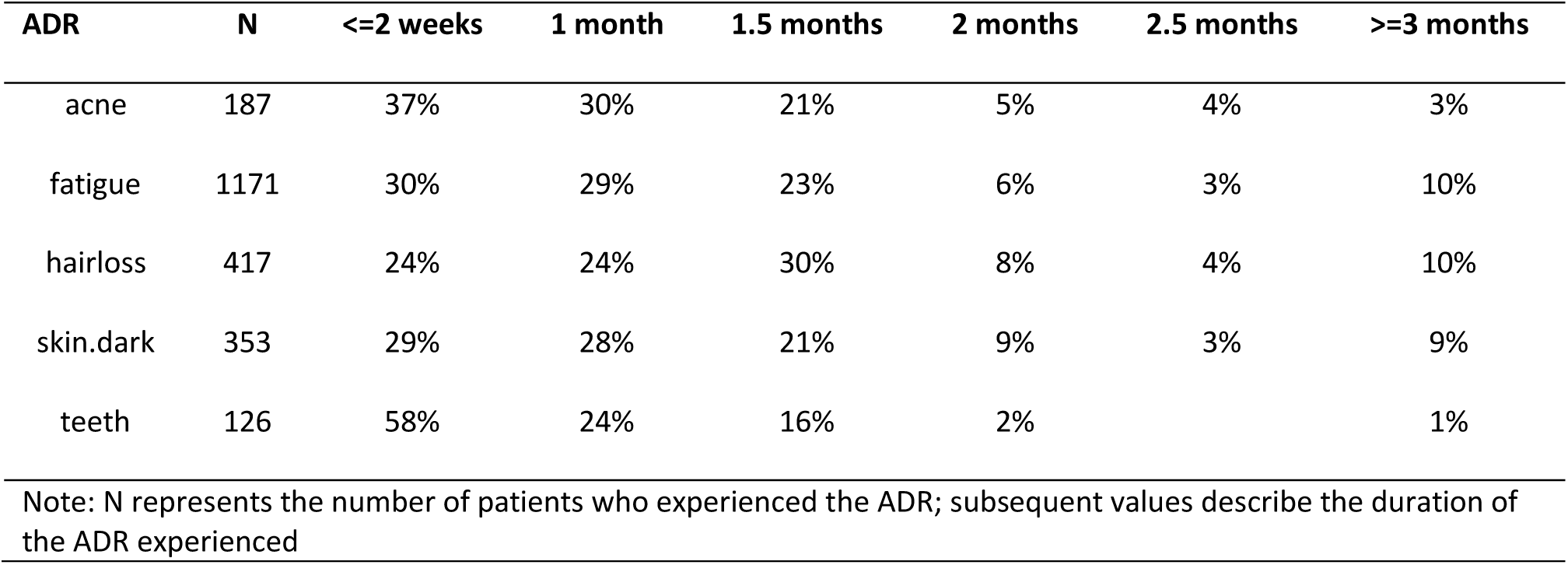
Duration of passive ADRs; N = 1333.

#### ADR Management by patients

Majority of the patients experiencing ADRs (n=1,827) are observed to seek resolution from their primary TB-treating HCP (71%), with 47% reaching out to a TC instead, or in addition to the former [Table 10]. While patients typically reach out to their treating HCP for an active ADR (66%), alternative doctors are a popular choice for passive ADRs (13%) [Table 11]. Those with an active ADR are more likely to seek a resolution (79%), than those with a passive ADR (72%). Females are generally more likely to consult their HCP or TC [Table 10; Tables S6 & S7, S1 Appendix]. Elderly are significantly less likely to consult with a TC, relative to other categories – for both active and passive ADRs. The site of TB does not have any bearing on ADR management.

**Table 10:** Action taken by patients for ADRs; summarized by management options; N = 1827 patients.

| Action taken | Females | Males | Total |
| --- | --- | --- | --- |
| consulted the doctor | 638 (74%) | 658 (68%) | 1296 (71%) |
| consulted TC | 441 (51%) | 422 (44%) | 863 (47%) |
| consulted another doctor (only valid for passive ADRs) | 89 (13%) | 84 (12%) | 173 (13%) |
| did nothing | 247 (29%) | 301 (31%) | 548 (30%) |

**Note:** a) Numbers are reported only for patients who did report ADRs; a total of 1,827 patients reported at least one ADR; b) Shares calculated for consulting another doctor are relative to the total patients who experienced a passive ADR, as the question was only posed to patients who reported a passive ADR

**Table 11:**
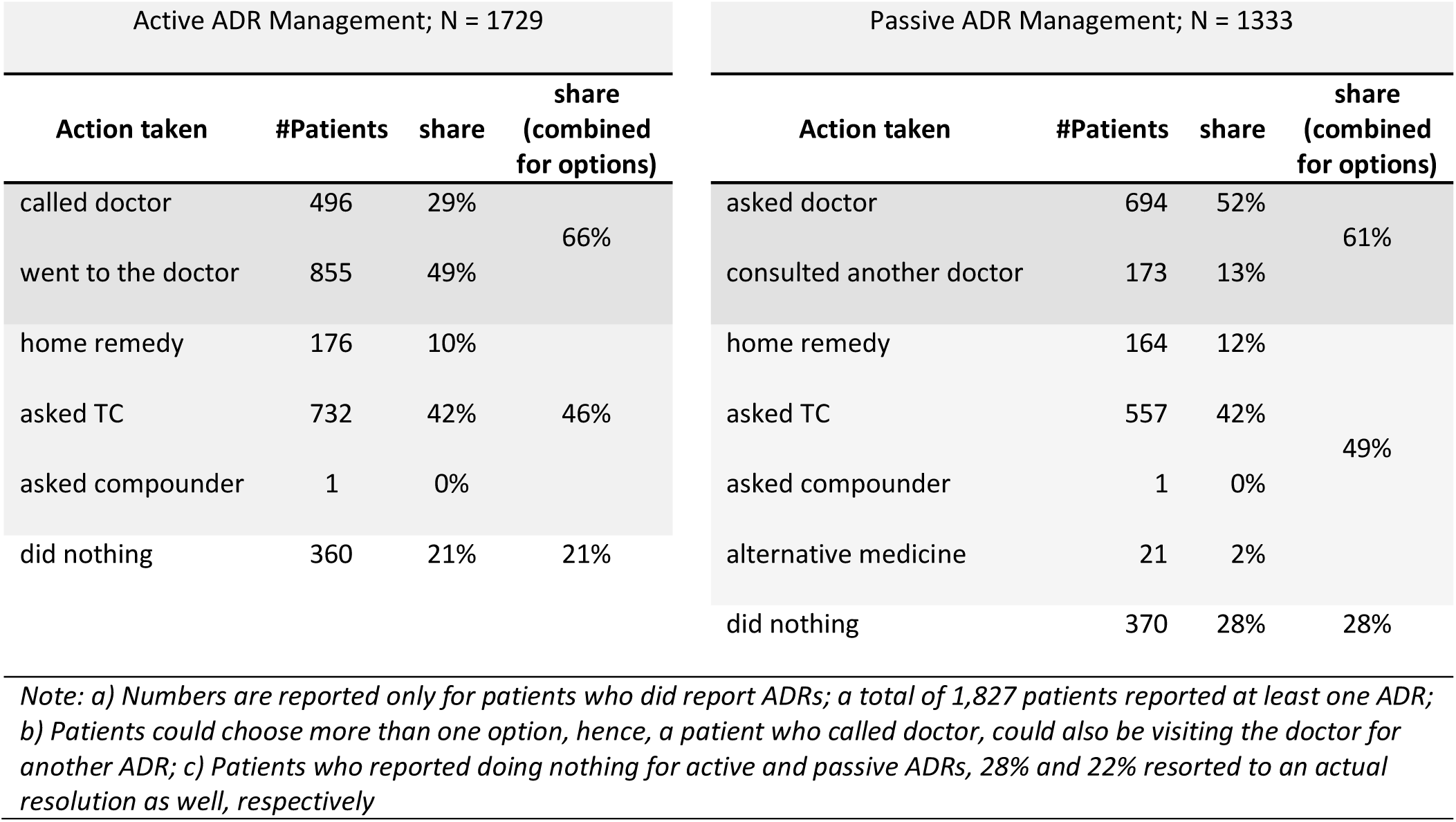
Action taken by patients for ADRs; summarized by ADR classification; N = 1827 patient.

#### Impact of ADRs on Treatment Outcomes

Treatment outcomes were available for 1,927 patients at the time of the study, classified as: 1) completed, 2) cured, 3) died, 4) failure, and 5) lost to follow up. Of these, completed and cured were considered successful [Table 12]. The total number of ADRs showed a significant negative association with treatment outcomes: each additional ADR reduces the odds a successful treatment outcome by 12% (OR = 0.88, p = 0.024) [Table 13]. For example, if the baseline probability of success was 70% (Odds = 0.7/0.3 = 2.33), experiencing two ADRs would reduce the probability of success to 64.4% (Odds = 2.33*0.88*0.88 = 1.8). If the baseline probability was instead higher (90%), the probability of success would decline to about 87.5%. While the decline appears modest, its impact at the population level could be considerable: in a cohort of 100,000 patients with an initial 90% probability of success, this reduction would translate into approximately 2,500 fewer patients cured (from 90,000 to ~87,500).

**Table 12:** Treatment outcomes of patients assessed in the study.

| Outcome | #N | Considered |
| --- | --- | --- |
| cured | 14 (0.7%) | Yes |
| died | 35 (1.6%) | Yes |
| lost to follow up | 12 (0.6%) | Yes |
| not evaluated | 2 (<0.1%) | No |
| pending | 159 (7.5%) | No |
| transferred out | 16 (0.8%) | No |
| treatment complete | 1,862 (88%) | Yes |
| treatment failure | 4 (0.2%) | Yes |
| treatment regimen changed | 20 (0.9%) | No |

**Table 13:**
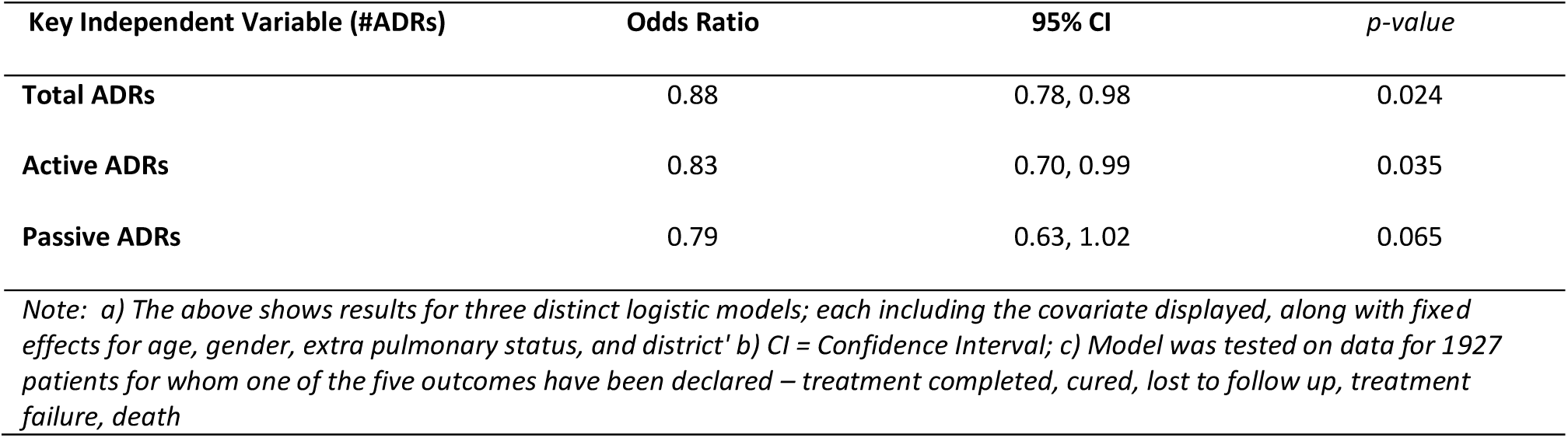
Results from logistic regression model(s) with binary treatment outcome as dependent variable.

| Key Independent Variable (#ADRs) | Odds Ratio | 95% CI | p-value |
| --- | --- | --- | --- |
| Total ADRs | 0.88 | 0.78, 0.98 | 0.024 |
| Active ADRs | 0.83 | 0.70, 0.99 | 0.035 |
| Passive ADRs | 0.79 | 0.63, 1.02 | 0.065 |

*Note: a) The above shows results for three distinct logistic models; each including the covariate displayed, along with fixed effects for age, gender, extra pulmonary status, and district' b) CI = Confidence Interval; c) Model was tested on data for 1927 patients for whom one of the five outcomes have been declared – treatment completed, cured, lost to follow up, treatment failure, death*

When fitted separately, an active ADR is found to reduce odds of a successful outcome by 17% (p = 0.035), while the corresponding figure for passive ADR is 21%, which is not significant (p = 0.065). This suggests that active ADRs are more consistent with compromising the completion of treatment, whereas passive ADRs may have a more variable impact across population. Among individual ADRs, vomiting was associated with the greatest impact, reducing the odds of a successful outcome by 61% (OR = 0.39; p = 0.006) [Table 14]. No other single ADR showed a significant association, indicating that the cumulative burden of multiple ADRs is generally more important in affecting outcomes than any single reaction, apart from vomiting.

**Table 14:** Results from logistic regression model with binary treatment outcome as dependent variable.

| Key Independent Variable (ADR) | Odds Ratio | 95% CI | p-value |
| --- | --- | --- | --- |
| diarrhoea | 1.03 | 0.41, 2.97 | >0.9 |
| drug fever | 1.08 | 0.54, 2.21 | 0.8 |
| itching | 0.81 | 0.40, 1.70 | 0.6 |
| joint pains | 0.74 | 0.38, 1.44 | 0.4 |
| trepidation | 1.54 | 0.77, 3.09 | 0.2 |
| vomit | 0.39* | 0.20, 0.76 | 0.006 |
| acne | 0.55 | 0.19, 1.69 | 0.3 |
| fatigue | 1.26 | 0.63, 2.51 | 0.5 |
| hair loss | 1.23 | 0.50, 3.30 | 0.7 |
| skin darkening | 0.57 | 0.25, 1.39 | 0.2 |
| teeth blackening | 1.06 | 0.36, 3.71 | >0.9 |

*Note: a) The above shows results for a single logistic model with binary independent variables for each of the 11 ADRs probed in the survey, along with fixed effects for age, gender, extra pulmonary status, and district' b) CI = Confidence Interval; c) Model was tested on data for 1927 patients for whom one of the five outcomes have been declared – treatment completed, cured, lost to follow up, treatment failure, death*

## 5. Discussion

To the best of our knowledge, this is the first study globally, to document ADR patterns among TB patients, using a patient-centric approach, across diverse socio-demographic categories, in a stratified and adequately powered sample, and spanning a large geographical region. Through the holistic assessment of incidence, duration and resolutions associated with ADRs, the study presents a transparent and nuanced insight into how patients approach long term treatment. It also introduces a novel ADR taxonomy, presents the specific risk propensities of different patient profiles, and illustrates the resulting impact on treatment outcomes. This constitutes critical evidence that can inform patient management protocols for TB, as well as for other chronic diseases where medication adherence is critical.

### Incidence & Barriers to reporting

Research on ADR incidence among DS-TB patients continues to be scarce [36]. While a meta-analysis found ADR incidence to vary between 8.4% and 83.5% [20], such measures include those on SLDs. For FLDs, the prevalence of ADRs is estimated to vary from 2.3% to 29% in various Indian studies [18,20,37], a finding much lower than what we report (86%). While these findings are highly dependent on mode of reporting (digital, telephonic, in-person), methods (clinical or self-reported), and reporting systems [38], under-reporting of ADRs is common in developing countries [22, 39]. Considering the significant likelihood of sub-standard or counterfeit pharmaceutical products in the latter [28], ADR reporting is crucial for factors beyond just adherence. Moreover, lack of reporting reflects and affects the priority given to ADRs experienced by patients – a vicious cycle propagated by lack of patient-HCP communication [39], with a serious impact on treatment completion rates [40–42]. Additionally, available reporting methods and guidelines are considered complicated and inadequate by healthcare workers [38]. However, as evidence suggests, if reported and proactively managed, the larger number of ADRs can actually improve outcomes, through better patient monitoring [43].

### Lack of support by healthcare providers & treatment coordinators

The present situation, however, requires much to be done. Earlier literature, and findings from our study – note the dismissive attitudes adopted by HCPs towards ADRs [39,44]. The qualitative interviews revealed the lack of resolutions available to patients inspite of reaching out to their HCPs and TCs, while the quantitative findings find a small but significant share of patients relying on alternative HCPs. Not everyone, however, can afford or access an alternative practitioner [45], exacerbating the risk posed.

### Significance of Passive ADRs

The risk is increased for passive ADRs, a category of ADRs not well-recognized in available NTEP documentation pertaining to ADR management [35], or in other meta-studies which detail symptom-based approaches to ADR management [36]. This is because earlier literature has typically focused on what we have classified as active ADRs: gastro-intestinal disorders (gastritis, etc.), dermatological reactions, nervous system challenges (dizziness, peripheral neuropathy, etc.)., arthralgia (joint pains), and hepatoxicity [20,40,46,47].

Our analyses, however, finds passive ADRs to be a significant risk-factor, owing ironically to their mild nature, albeit persisting over longer durations. Their occurrence typically coincides with the period when TB symptoms have subsided, increasing the likelihood that patients perceive treatment as unnecessary. This increases the likelihood of treatment discontinuation, which in turn is strongly linked to relapse and the emergence of drug-resistant TB [48]. Moreover, even though our econometric results estimated a weaker (p = 0.065) association between passive ADRs and treatment outcomes, the larger effect size (21% lower odds of successful treatment outcome with each additional passive ADR, relative to 17% for active ADRs) combined with the longer duration of these ADRs [Table 9] highlight their clinical relevance. Importantly, passive ADRs may be more amenable to timely counselling and psychosocial support, presenting an opportunity for programmatic action within NTEP and similar national TB programs.

### Patient and provider perceptions of ADRs

While our study did not directly assess the perceived severity of ADRs experienced, a lack of agreement between patients and HCPs is evident considering the external resolutions sought by patients. Prior evidence indicates that patients may discontinue treatment if they perceive an ADR to be severe, even when HCPs think and communicate otherwise [39]. Strengthening patient-HCP communication, and fostering an acceptance mindset around patients’ perception of ADRs among HCPs, even and especially for passive ADRs, is hence critical to reduce risk. Social support and information seeking are commonly identified patient strategies in literature, with timely guidance linked to better adherence [49].

### Differentiated care – patient risk profiling

Providing identical, blanket counselling to all patients - particularly through proactive approaches that are offered irrespective of individual risk or need - is time-intensive, inefficient, and unrealistic in resource-constrained, high-burden settings. By validating ADR patterns across individual patient profiles, our study unveiled specific risk-groups, which can be used for differentiated counselling [Table 15]. Gender and age-related differences were particularly salient, with women experiencing more ADRs, often persisting for longer durations, while elderly patients were more prone to specific ADR types, as is also evidenced in literature [50–52].

**Table 15:** Counselling Prioritization Matrix for ADR Management by Patient Profile.

| Patient Group | Priority Active ADRs | Priority Passive ADRs | Comorbidities | Communicate ADRs & seek resolution | Self-care & rest | Reassure: skin & hair loss temporary | Continue adherence | Guidance for vomiting* | Overall Guidance |
| --- | --- | --- | --- | --- | --- | --- | --- | --- | --- |
| Women (employed / unemployed) | Drug Fever, Vomit (high-risk), Trepidation | Skin/hair changes, fatigue |  |  | ✓ | ✓ | ✓ | ✓✓ | Emphasize self-care; reassure that visible ADRs are temporary. |
| Employed males |  | Low reporting of minor ADRs |  | ✓ |  |  | ✓ | ✓ | Encourage proactive ADR reporting. |
| Younger patients | Vomiting (high-risk) | Cutaneous ADRs (acne, hair loss, skin darkening) |  | ✓ | ✓ | ✓ | ✓ | ✓✓ | Reinforce adherence; address stigma. |
| Elderly patients | Gastro-intestinal (diarrhoea, vomit), joint pains | Few cutaneous ADRs reported | ✓ |  |  |  | ✓ | ✓ | Prioritize comorbidities and ADR-related risks. |
| Notes:<br>1) All counselling aspects are relevant for all patients, but additional emphasis ("✓") is suggested for groups at higher risk.<br>2) Active ADRs generally require stronger clinical management (e.g., vomiting in younger patients, gastrointestinal and joint pain in elderly), while passive ADRs call for counselling and psychosocial support (e.g., skin/hair changes in women and younger patients, under-reporting in men). The "✓" markers in the table indicate where emphasis should be placed for each group, depending on whether ADRs are active or passive.<br>3) For vomit - two "✓✓" highlight that the said risk group is more likely than others to experience the same. It is highlighted for all as it is independently predicted to reduce odds of successful treatment completion |  |  |  |  |  |  |  |  |  |

Gender-based differences in drug response and ADR occurrence are well established, attributed to pharmacokinetic, immunological, and hormonal factors [53–55]. TB-specific studies have similarly documented earlier onset and greater prevalence of ADRs among women [12,14,56]. In our study, over 76% of women reported ≥2 ADRs, compared with 66% of men. This higher reporting may also relate to heightened stigma around visible ADRs: 30% of women reported hair loss (vs. 11% of men) and 19% reported skin darkening (vs. 15% of men). Women are also generally more prone to drug-induced risks [50, 55], and a significantly higher share (41% vs 29%) of them experience vomiting – the only identified ADR in our analyses to independently affect treatment outcomes [Table 14]. The challenge extends to men in a different way: socio-cultural norms of masculinity often equate sharing an illness or seeking care for the same with weakness [58,59]. This discourages care-seeking, ADR reporting, and resolution-seeking [58,59], ultimately contributing to poorer treatment adherence [60].

Among elderly, the risk is heightened due to physiological changes in drug metabolism, as well as comorbidities which increase the risk of drug interactions and cumulative toxicity [51,52,61]. Advanced age has been evidenced as an independent predictor of severe ADRs, even after adjusting for sex and comorbidities [62,63]. In our study, elderly patients were significantly more likely to experience diarrhoea and musculoskeletal ADRs, though without reporting longer durations for these. Conversely, lower reporting of cutaneous ADRs (e.g., skin darkening, hair loss) may reflect normalization of such symptoms, competing health priorities, or their general de-prioritization. Elderly are also significantly less likely to reach out to TCs (31%), relative to younger patients (42% - 45%). These patterns underscore the importance of proactive communication by TCs, which could accelerate resolution of milder ADRs and mitigate associated risks. Younger patients, while otherwise resilient, report a significantly high prevalence of vomiting (41%), a result which is identically found for women, and is estimated to independently reduce the odds of treatment completion by 61%. Additionally, while the higher reporting of cutaneous ADRs among this group may not be perceived as clinically severe, this class of ADRs often carries considerable psychosocial weight. When compounded by the stigma associated with TB itself, this can further undermine adherence if not addressed through timely counselling and support.

### Recommendations

While ADR management is critical to the success of long-term treatment regimens, current program protocols provide limited guidance, often leaving patients to navigate the resolution pathway on their own. We recommend shifting from the presently followed symptom-based approach [36] to a more proactive, patient-centred ADR management strategy, which takes into account the dual ADR taxonomy. Five key recommendations are detailed, in order to augment the broader ADR management strategy within NTEP.

#### Risk Profiling at Treatment Initiation

Profiling newly diagnosed patients based on identified propensities to active and passive ADRs, in addition to symptoms and other pre-existing factors, can enable proactive and tailored counselling.

#### Integration of ADR monitoring into Nikshay

Embedding structured ADR reporting using closed-ended (Yes/No) responses for all identified ADRs, including passive ones, within the government TB data system would enhance reporting accuracy, generate more disaggregated insights across socio-demographic groups, and enable timely support for at-risk patients. The dual taxonomy offers a pragmatic way to tailor counselling. For instance, persistent ADRs such as fatigue or skin darkening may be addressed through self-management strategies, including nutrition support and simple home remedies, while active ADRs such as vomiting or joint pains could be flagged for clinical intervention.

#### Strengthen TC Training & Customized patient assignments

Dedicated training, in order to deliver *time-based, risk-specific counselling* can help TCs to optimize their efforts towards patient counselling. For example, focusing on active ADRs during treatment initiation and introducing information on passive ADRs in the second month can reduce information overload, improving patient comprehension and recall. Further, prioritizing counselling by risk-groupings can help address concerns more effectively for relevant groups. This could mean encouraging men to proactively report ADRs early in treatment, or addressing cutaneous ADR concerns with younger patients before they experience them. Efficiency could further be improved by aligning TCs to specific patient groups over defined periods, allowing them to deliver more consistent and targeted counselling. The ADR collateral developed for JEET TCs, which outlines resolution pathways across a four-grade severity matrix, can serve as a practical training and implementation tool for this purpose [Supp Text S4, S5], together with the counselling prioritization matrix (Table 15).

#### HCP Sensitization

Training and sensitization modules for HCPs should reinforce the importance of taking patient complaints seriously, strengthening patient-HCP communication, and ensuring clear referral pathways to alternative providers or specialists when necessary. This is especially true for passive ADRs where the common tendency to overlook passive ADRs can undermine adherence among patients.

#### Structured Patient Education

Illustrated guides and counselling aids can help equip patients and caregivers to recognize ADRs, self-manage minor ones, and seek timely help for more severe reactions.

Taken together, these patient-centric approaches, if embedded within national TB protocols, can help bring ADR management to the forefront of patient counselling, easing the burdens often shouldered by patients and their caregivers alone. We encourage program staff in India and other high-burden settings to adapt the findings of our study, particularly the active - passive ADR framework and demographic risk-groupings, within their program protocols. The ADR collateral developed and used by JEET TCs, included in the supplementary materials, already incorporates passive ADRs and provides a strong foundation. Building on this existing collateral by incorporating risk-groupings (Table 15) offers a practical and feasible pathway to deliver more differentiated and effective patient support.

## 6. Conclusion

This study demonstrates the near-universal burden of ADRs among TB patients and their impact on adherence and treatment outcomes. By introducing a patient-centred active-passive ADR taxonomy, we highlight how persistent, under-recognized passive ADRs can quietly undermine adherence, particularly in later treatment phases. Demographic disparities, validated across a powered and stratified sample, provide an evidence-based roadmap for differentiated, gender- and age-responsive counselling. Augmenting existing ADR management protocols with the identified active-passive framework and risk based counselling within NTEP frameworks can help arrest treatment interruptions and strengthen programmatic response. Further research on scalability and cost-effectiveness will be critical for informing financing and prioritization, but the evidence presented here demonstrates both the urgency and feasibility of embedding structured ADR management into TB care in India and other high-burden settings.

## 7. Future Research

These findings highlight several areas for further investigation. First, examining ADRs among patients with comorbidities, and how these are managed across different care conditions, could inform how differentiated counselling approaches may be adapted at scale. Second, as ADR reporting becomes integrated within government databases, including through Nikshay Sampark and other pharmacovigilance mechanisms, their linkage with treatment outcomes could be revisited across a larger population base. Third, an assessment of ADRs in more controlled prescription settings, which can help assess differential incidence between those on FDCs or not, can help aid understanding in managing them. Lastly, evaluating the cost-effectiveness of various ADR management strategies, particularly in relation to treatment success rates and the opportunity costs of relapse or drug-resistant TB, could help justify greater investment in ensuring treatment continuity, a critical requirement for reducing and ultimately eliminating TB.

## 8. Limitations

Our analysis has certain limitations which warrant attention. First, the quantitative phase of our analysis primarily relies on closed-ended questions to enquire about the ADRs so experienced. This design was chosen to minimize recall bias, as patients could select from a standardized list rather than relying solely on memory. To reduce the risk of omission, an open-ended follow-up question was also included to capture any additional ADRs reported by patients. Nonetheless, this approach may still lead to over- or under-reporting of certain ADRs. Notwithstanding this possible bias, the authors found no reason to suspect the same from the results so obtained, as the results obtained were consistent with prior literature (e.g., pulmonary vs extra pulmonary TB not influencing ADRs, women experiencing more ADRs). Second, the incidence of ADRs assessed in our study is self-reported, and not clinically validated. Our analysis does not attempt to understand the actual veracity or the underlying cause of the ADRs so experienced, which may very well be due to a different medication being taken by the patient, in addition to the ATT drugs. Third, as the quantitative survey was administered by patients’ own TCs, the possibility of social desirability bias cannot be fully excluded. To mitigate this, TCs received standardized training, the questionnaire was carefully piloted, and patients were assured that their responses would remain confidential and would not affect their treatment. At the same time, being interviewed by their own TCs may also have created an opposite effect - patients could feel more comfortable and willing to share ADRs openly, given the trust and familiarity with their TC. The consistently higher ADR incidence observed in our study compared to previous literature suggests that, in sum, this context likely encouraged more candid reporting, and any residual bias was minimal. Fourth, a small share of patients reported feeling abdomen pain, and weakness in eyesight as additional ADRs. Since these patients were much less in number, we did not report these figures. Further studies on this topic will benefit from including a more comprehensive list of ADRs observed. Fifth, we use a derived dichotomous outcome variable to understand the impact of ADRs on treatment outcomes. Here, unsuccessful outcomes include treatment failure, death, and lost to follow-up, and each of these outcomes may have their own risk profiles. All patients who had a treatment interruption greater than one month in duration are considered as being lost to follow-up. However, it cannot be determined if patients continued the treatment later, and if so, whether they were able to complete the treatment with a positive outcome. Hence, including lost to follow-up has the potential to bias these results. Some previous studies have not included lost to follow-up in their analyses for similar reasons [64]. However, we remain conservative and followed the baseline criteria of including patients who were under the active management of a TC, and had their outcomes reported at least a month after the date of diagnosis. Additionally, in our analysis, lost to follow-up makes up 0.6% of our dataset. Including lost to follow-up in analysis where these cases make less than <5% of the overall population generally leads to little bias [65]. Lastly, majority of patients reported treatment completion which was based on the provider declaring that patient need not take any more medications. Since cure rates are low due to lack of smear testing in the private sector, the metric of successful treatment completion itself has certain limitations.

## Supporting information

Supp 3 (Qual Guide)

## Data Availability

All of the data utilized is available in agithub repository: https://github.com/ridhimasodhi/ADR.TB

## Acknowledgments

We would like to thank Dr. Ashwani Khanna, a senior pulmonologist and former State TB Officer for New Delhi, for reviewing the manuscript and providing valuable edits, particularly on provider–patient communication and the importance of supporting patients experiencing passive ADRs.

## List of abbreviations

ADR: Adverse Drug Reaction
JEET: Joint Effort for Elimination of Tuberculosis
OLS: Ordinary least squares
CI: Confidence interval
IQR: Interquartile range
PPSA: Private Provider Support Agency
TB: Tuberculosis
NTEP: National Tuberculosis Elimination Program
NSP: National Strategic Plan for Elimination of Tuberculosis

## Supplementary Information – Legend

1. S1 Appendix: Additional tables and figures, covering sample size computations, definitions of different treatment outcomes, patient profiles, ADR incidence by age and regions, and information on ADR management by different covariates / groups
2. S2 Appendix: Discussion Guide for Qualitative Phase
3. S3 Appendix: Questionnaire for Quantitative Phase
4. S4 Appendix: ADR Management Guide for Treatment Supporters (English)
5. S5 Appendix: ADR Management Guide for Treatment Supporters (Hindi)

