## Supplementary material for "Adverse Drug Reactions in Tuberculosis Treatment: Incidence, Duration and Resolution Pathways from a Mixed-Methods Patient-centric Study in India": Supp 1 (Additional Tables & Figures)

**S1 Appendix**

### Adverse Drug Reactions in Tuberculosis Treatment: Incidence, Reporting and Outcomes; Insights from a Mixed-Methods Study Across Eight Indian Cities

***Authors**: Ridhima Sodhi[1¶*], Tunisha Kapoor[1&], Vindhya Vatsyayan[1&], Iti Seth[2&], Harsh Chandra[2&], Nishita Gill[2&], Manoj Singh[1&], Arnab Pal[1&], Shamim Mannan[1&]*

1. William J Clinton Foundation, New Delhi, India
2. Treemouse Research & Design Private Limited, New Delhi, India

#### 1. *Purposive Sampling Approach – Qualitative Phase*

Patients: A total of 40 patients were finally interviewed across three districts – Delhi, Jaipur, and Bhopal, with at least 10 patients from each site. Participants were selected basis socio-demographic characteristics (age, gender, stage of TB treatment, profession, and marital status), and stage of treatment (Fig 2). Three stages were recognized, a) the intensive phase or the first two months, b) the continuous phase or when the patients have completed at least 2 months of treatment, and c) post treatment completion. The selection prioritized patients who have faced more challenges during treatment, in order to further insights around resolution management.

Family Members: A total of 5 family members of patients were interviewed across the three sites, and questions elicited information on the type and extent of support provided by family members.

Treatment Coordinators**:** A total of 20 TCs were interviewed (Jaipur- 4, Delhi – 11, Bhopal - 5). The TCs were selected basis their performance – distributed across high performing (10), average/new and low-performing ones (11). The patients who were interviewed also included patients who were being guided by the TCs from these 2 categories. The feedbacks for TCs were drawn from program leaders.

Doctors: A total of 12 doctors were interviewed (Jaipur - 5, Delhi - 4, Bhopal - 3), with questions covering their perception of the JEET PPSA model, and patient management strategies.

Hub Agents: A total of 8 hub agents, who had worked with >=10 patients, were interviewed (Jaipur - 3, Delhi - 4 and Bhopal - 1).

Program Staff: Two program directors and two field staff were interviewed to offer insights around programmatic operations and provider engagement strategies.

#### 2. *Sample Size*

Since our variable of interest is the proportion of population, we used the following formula for estimating required sample size[1].

$$n = \frac{(z)^2\, p(1-p)}{E^2 + \ (z^2 * p * (1-p)/N)}$$

whereby,
n = calculated sample size,
N = underlying population
z = confidence level at the chosen probability level (1.96 for 95%),
p = anticipated response distribution (we assume a conservative maximum estimate of 50%), and
E = tolerable margin of error

We have segregated population and sample count as per intensive and continuous phase. We decide on their individual sample sizes by using proportional stratification method, wherein the sample size of each stratum or treatment phase is proportionate to the population size of that treatment phase or stratum. Specifically, we make use of the following equation:

[1] Sampling Design & Analysis, S. Lohr, 1999, Chapter 2, equation 2.17

$$n(i) = \frac{N(i) * n}{N}$$

n(i) = sample size of the intensive/continuous phase,
N(i) = patient base of the intensive/continuous phase,
N = total patient base
n = total sample size computed using prescribed C.I. and margin of error

#### 3. *Definitions of treatment outcomes for drug susceptible TB patients*

***S1 Table***
**Definitions of treatment outcomes for drug susceptible TB patients** [1]

| Treatment outcome | Definition | Study outcome | Considered for the study |
|---|---|---|---|
| Cured | Microbiologically confirmed TB patients at the beginning of treatment who was smear or culture negative at the end of the complete treatment | Successful treatment | Yes |
| Treatment complete | Completed treatment without evidence of failure or clinical deterioration but with no record that the smear or culture results of biological specimen in the last month of treatment was negative | | |
| Lost to follow up | Treatment was interrupted for one consecutive month or more | Unsuccessful treatment | |
| Died | Died during the course of anti-TB treatment | | |
| Treatment failure | Biological specimen is positive by smear or culture at end of treatment | | |
| Not evaluated | Patients for whom no treatment outcome is assigned; also includes former transfer outs | Other outcomes (not considered) | No |
| Treatment regimen changed | A TB patient who is on first line regimen and has been diagnosed as having DR TB and switched to drug resistant TB regimen prior to being declared as failed | | |
| Wrongly diagnosed | A patient who is wrongly diagnosed of TB | | |
| Transferred | A patient who has transferred to another facility or state, prior to the outcome being declared | | |

#### 4. *Demographic characteristics of distinct patient categories*

***S2 Table***
*Demographic characteristics of distinct patient categories; N = 2124 patients*

| Characteristic | dependents N = 440 | elderly dependents N = 216 | employed females N = 144 | employed males N = 748 | homemaker N = 491 | other N = 85 | p-value |
|---|---|---|---|---|---|---|---|
| **Males** | 230 (52%) | 125 (58%) | 0 (0%) | 748 (100%) | 0 (0%) | 58 (68%) | <0.001 |
| **Age Category** | | | | | | | <0.001 |
| 1. <=22 | 325 (74%) | 0 (0%) | 39 (27%) | 87 (12%) | 64 (13%) | 26 (31%) | |
| 2. 23-59 | 115 (26%) | 0 (0%) | 100 (69%) | 604 (81%) | 427 (87%) | 59 (69%) | |
| 3. >=60 | 0 (0%) | 216 (100%) | 5 (3.5%) | 57 (7.6%) | 0 (0%) | 0 (0%) | |
| **Age** | | | | | | | <0.001 |
| Median (IQR) | 20 (18, 23) | 65 (62, 72) | 28 (22, 35) | 37 (27, 50) | 30 (25, 43) | 33 (22, 48) | |
| Mean | 23 | 68 | 30 | 39 | 34 | 34 | |
| SD | 10 | 7 | 11 | 13 | 11 | 13 | |
| **Extra Pulmonary** | 141 (32%) | 69 (32%) | 56 (39%) | 191 (26%) | 182 (37%) | 19 (22%) | <0.001 |
| **District** | | | | | | | 0.077 |
| ahmedabad | 62 (14%) | 56 (26%) | 21 (15%) | 136 (18%) | 110 (22%) | 18 (21%) | |
| bhopal | 31 (7.0%) | 21 (9.7%) | 26 (18%) | 67 (9.0%) | 39 (7.9%) | 8 (9.4%) | |
| darbhanga | 50 (11%) | 29 (13%) | 4 (2.8%) | 109 (15%) | 56 (11%) | 11 (13%) | |
| delhi | 88 (20%) | 24 (11%) | 28 (19%) | 82 (11%) | 54 (11%) | 7 (8.2%) | |
| gurgaon | 44 (10%) | 10 (4.6%) | 16 (11%) | 73 (9.8%) | 39 (7.9%) | 5 (5.9%) | |
| indore | 45 (10%) | 20 (9.3%) | 18 (12%) | 83 (11%) | 64 (13%) | 9 (11%) | |
| jaipur | 64 (15%) | 39 (18%) | 10 (6.9%) | 87 (12%) | 77 (16%) | 16 (19%) | |
| surat | 56 (13%) | 17 (7.9%) | 21 (15%) | 111 (15%) | 52 (11%) | 11 (13%) | |

Note: p values are given for Kruskal-Wallis rank sum test

#### 5. *Age related risk factors for individual ADRs*

***S3 Table***

*Tukey Tests; Significance of difference in means for incidence of distinct ADRs reported among distinct age groups; N = 2124 patients*

| | **Age Group Pair** | *Adjusted P (Sidak correction)* |
|---|---|---|
| | 23-59 &<=22 years | 0.894 |
| **diarrhoea** | >=60 &<=22 years | 0.0002* |
| | >=60 & 23-59 years | 0.0001* |
| | 23-59 &<=22 years | 0.069 |
| **drug fever** | >=60 &<=22 years | 0.053 |
| | >=60 & 23-59 years | 0.650 |
| | 23-59 &<=22 years | 0.085 |
| **itching** | >=60 &<=22 years | 0.277 |
| | >=60 & 23-59 years | 0.998 |
| | 23-59 &<=22 years | 0.0011* |
| **joint pains** | >=60 &<=22 years | 0.0143* |
| | >=60 & 23-59 years | 0.928 |
| **trepidation** | 23-59 &<=22 years | 0.738 |

| | | |
|---|---|---|
| | >=60 &<=22 years | 0.542 |
| | >=60 & 23-59 years | 0.818 |
| | 23-59 &<=22 years | 0.0003* |
| **vomit** | >=60 &<=22 years | 0.068 |
| | >=60 & 23-59 years | 0.845 |
| | 23-59 &<=22 years | 0.000* |
| **acne** | >=60 &<=22 years | 0.000* |
| | >=60 & 23-59 years | 0.184 |
| | 23-59 &<=22 years | 0.577 |
| **fatigue** | >=60 &<=22 years | 0.226 |
| | >=60 & 23-59 years | 0.535 |
| | 23-59 &<=22 years | 0.000* |
| **hair loss** | >=60 &<=22 years | 0.000* |
| | >=60 & 23-59 years | 0.0365* |
| | 23-59 &<=22 years | 0.0067* |
| **skin darkening** | >=60 &<=22 years | 0.007* |
| | >=60 & 23-59 years | 0.557 |
| | 23-59 &<=22 years | 0.957 |
| **teeth darkening** | >=60 &<=22 years | 0.0437* |
| | >=60 & 23-59 years | 0.0372* |

Note: a) p value gives the results from Tukey test to test the significance of difference in ADR incidence for distinct age groups; b) * or a significant value shows that the paired age-groups are statistically significantly different from each other with a p value of lower than 0.05

#### 6. *Duration of ADRs*

***S4 Table***

*Summary of duration of active ADRs; N = 1729 (those who experienced at least one active ADR)*

| | Age Class | | | | Gender | | | Categories | | | | | | | Site of Disease | | |
|---|---|---|---|---|---|---|---|---|---|---|---|---|---|---|---|---|---|
| | **<=22** | **23-59** | **>=60** | *p-value* | **female** | **male** | *p-value* | **dependents** | **elderly dependents** | **employed females** | **employed males** | **homemaker** | **other** | *p-value* | **pulmonary** | **extra pulmonary** | *p-value* |
| **#Patients** | **444** | **1063** | **222** | | **822** | **907** | | **347** | **174** | **126** | **584** | **426** | **72** | | **1198** | **531** | |
| **diarrhoea** | | | | 0.9 | | | 0.3 | | | | | | | 0.7 | | | <0.001 |
| <=1 week | 17 (45%) | 53 (53%) | 23 (55%) | | 38 (48%) | 55 (54%) | | 17 (55%) | 14 (47%) | 7 (64%) | 34 (52%) | 16 (44%) | 5 (71%) | | 83 (61%) | 10 (22%) | |
| 2-3 weeks | 11 (29%) | 20 (20%) | 7 (17%) | | 15 (19%) | 23 (23%) | | 8 (26%) | 5 (17%) | 0 (0%) | 15 (23%) | 9 (25%) | 1 (14%) | | 25 (19%) | 13 (29%) | |
| 1 month | 9 (24%) | 23 (23%) | 9 (21%) | | 23 (29%) | 18 (18%) | | 6 (19%) | 8 (27%) | 4 (36%) | 14 (22%) | 9 (25%) | 0 (0%) | | 22 (16%) | 19 (42%) | |
| 1.5 months | 0 (0%) | 3 (3.0%) | 2 (4.8%) | | 2 (2.5%) | 3 (3.0%) | | 0 (0%) | 2 (6.7%) | 0 (0%) | 1 (1.5%) | 2 (5.6%) | 0 (0%) | | 3 (2.2%) | 2 (4.4%) | |
| 2 months | 0 (0%) | 1 (1.0%) | 0 (0%) | | 0 (0%) | 1 (1.0%) | | 0 (0%) | 0 (0%) | 0 (0%) | 0 (0%) | 0 (0%) | 1 (14%) | | 1 (0.7%) | 0 (0%) | |
| >=2 months | 1 (2.6%) | 0 (0%) | 1 (2.4%) | | 1 (1.3%) | 1 (1.0%) | | 0 (0%) | 1 (3.3%) | 0 (0%) | 1 (1.5%) | 0 (0%) | 0 (0%) | | 1 (0.7%) | 1 (2.2%) | |
| **drug fever** | | | | 0.5 | | | 0.13 | | | | | | | 0.5 | | | 0.15 |
| <=1 week | 74 (31%) | 173 (34%) | 37 (37%) | | 129 (32%) | 155 (36%) | | 57 (32%) | 27 (36%) | 25 (34%) | 97 (35%) | 61 (31%) | 17 (46%) | | 208 (34%) | 76 (33%) | |
| 2-3 weeks | 58 (24%) | 131 (26%) | 24 (24%) | | 96 (24%) | 117 (27%) | | 49 (28%) | 18 (24%) | 14 (19%) | 82 (30%) | 45 (23%) | 5 (14%) | | 161 (26%) | 52 (23%) | |
| 1 month | 54 (23%) | 88 (18%) | 19 (19%) | | 91 (22%) | 70 (16%) | | 33 (19%) | 17 (22%) | 16 (22%) | 41 (15%) | 46 (23%) | 8 (22%) | | 121 (20%) | 40 (17%) | |
| 1.5 months | 13 (5.5%) | 17 (3.4%) | 3 (3.0%) | | 18 (4.4%) | 15 (3.5%) | | 11 (6.2%) | 2 (2.6%) | 3 (4.1%) | 8 (2.9%) | 8 (4.0%) | 1 (2.7%) | | 25 (4.1%) | 8 (3.5%) | |

| | <=22 | 23-59 | >=60 | p-value | female | male | p-value | dependents | elderly dependents | employed females | employed males | homemaker | other | p-value | pulmonary | extra pulmonary | p-value |
|---|---|---|---|---|---|---|---|---|---|---|---|---|---|---|---|---|---|
| 2 months | 9 (3.8%) | 37 (7.4%) | 5 (5.1%) | | 27 (6.7%) | 24 (5.5%) | | 2 (1.1%) | 4 (5.3%) | 8 (11%) | 15 (5.4%) | 18 (9.0%) | 4 (11%) | | 32 (5.3%) | 19 (8.3%) | |
| >=2 months | 30 (13%) | 56 (11%) | 11 (11%) | | 45 (11%) | 52 (12%) | | 25 (14%) | 8 (11%) | 8 (11%) | 33 (12%) | 21 (11%) | 2 (5.4%) | | 62 (10%) | 35 (15%) | |
| **itching** | | | | 0.8 | | | 0.6 | | | | | | | 0.2 | | | 0.8 |
| <=1 week | 69 (44%) | 126 (40%) | 29 (43%) | | 108 (39%) | 116 (44%) | | 46 (41%) | 20 (38%) | 25 (57%) | 75 (43%) | 52 (37%) | 6 (38%) | | 148 (42%) | 76 (41%) | |
| 2-3 weeks | 32 (20%) | 88 (28%) | 20 (30%) | | 74 (27%) | 66 (25%) | | 25 (22%) | 16 (31%) | 10 (23%) | 44 (25%) | 42 (30%) | 3 (19%) | | 92 (26%) | 48 (26%) | |
| 1 month | 33 (21%) | 58 (18%) | 8 (12%) | | 58 (21%) | 41 (15%) | | 23 (20%) | 7 (13%) | 7 (16%) | 29 (17%) | 30 (21%) | 3 (19%) | | 68 (19%) | 31 (17%) | |
| 1.5 months | 10 (6.4%) | 10 (3.2%) | 3 (4.5%) | | 14 (5.1%) | 9 (3.4%) | | 9 (8.0%) | 3 (5.8%) | 0 (0%) | 5 (2.9%) | 5 (3.5%) | 1 (6.2%) | | 15 (4.2%) | 8 (4.3%) | |
| 2 months | 4 (2.5%) | 12 (3.8%) | 3 (4.5%) | | 10 (3.6%) | 9 (3.4%) | | 1 (0.9%) | 3 (5.8%) | 1 (2.3%) | 6 (3.4%) | 7 (4.9%) | 1 (6.2%) | | 11 (3.1%) | 8 (4.3%) | |
| >=2 months | 9 (5.7%) | 23 (7.3%) | 4 (6.0%) | | 11 (4.0%) | 25 (9.4%) | | 9 (8.0%) | 3 (5.8%) | 1 (2.3%) | 15 (8.6%) | 6 (4.2%) | 2 (12%) | | 22 (6.2%) | 14 (7.6%) | |
| **joint pain** | | | | >0.9 | | | 0.092 | | | | | | | 0.2 | | | 0.4 |
| <=1 week | 59 (28%) | 198 (31%) | 43 (31%) | | 135 (28%) | 165 (33%) | | 50 (31%) | 33 (31%) | 17 (24%) | 114 (33%) | 70 (27%) | 16 (40%) | | 203 (30%) | 97 (32%) | |
| 2-3 weeks | 61 (29%) | 150 (24%) | 28 (20%) | | 111 (23%) | 128 (25%) | | 40 (25%) | 24 (22%) | 24 (33%) | 86 (25%) | 55 (21%) | 10 (25%) | | 166 (25%) | 73 (24%) | |
| 1 month | 50 (23%) | 143 (23%) | 36 (26%) | | 125 (26%) | 104 (21%) | | 41 (25%) | 26 (24%) | 16 (22%) | 72 (21%) | 71 (27%) | 3 (7.5%) | | 149 (22%) | 80 (26%) | |
| 1.5 months | 16 (7.5%) | 26 (4.1%) | 5 (3.6%) | | 21 (4.4%) | 26 (5.1%) | | 8 (5.0%) | 4 (3.7%) | 2 (2.8%) | 16 (4.7%) | 13 (5.0%) | 4 (10%) | | 37 (5.5%) | 10 (3.3%) | |
| 2 months | 14 (6.6%) | 45 (7.1%) | 10 (7.2%) | | 38 (7.9%) | 31 (6.1%) | | 9 (5.6%) | 6 (5.6%) | 4 (5.6%) | 21 (6.2%) | 25 (9.6%) | 4 (10%) | | 50 (7.4%) | 19 (6.2%) | |
| >=2 months | 13 (6.1%) | 70 (11%) | 16 (12%) | | 48 (10%) | 51 (10%) | | 13 (8.1%) | 15 (14%) | 9 (12%) | 32 (9.4%) | 27 (10%) | 3 (7.5%) | | 71 (11%) | 28 (9.1%) | |
| **trepidation** | | | | 0.2 | | | 0.6 | | | | | | | 0.6 | | | 0.5 |
| <=1 week | 122 (46%) | 325 (49%) | 62 (42%) | | 270 (48%) | 239 (47%) | | 89 (45%) | 49 (43%) | 42 (52%) | 160 (48%) | 141 (46%) | 28 (60%) | | 350 (47%) | 159 (48%) | |
| 2-3 weeks | 63 (24%) | 172 (26%) | 43 (29%) | | 148 (26%) | 130 (25%) | | 54 (27%) | 30 (26%) | 18 (22%) | 87 (26%) | 86 (28%) | 3 (6.4%) | | 187 (25%) | 91 (27%) | |
| 1 month | 48 (18%) | 101 (15%) | 19 (13%) | | 88 (16%) | 80 (16%) | | 35 (18%) | 16 (14%) | 11 (14%) | 48 (15%) | 49 (16%) | 9 (19%) | | 118 (16%) | 50 (15%) | |
| 1.5 months | 13 (4.9%) | 26 (3.9%) | 9 (6.1%) | | 19 (3.4%) | 29 (5.7%) | | 9 (4.5%) | 8 (7.0%) | 0 (0%) | 19 (5.8%) | 9 (3.0%) | 3 (6.4%) | | 39 (5.2%) | 9 (2.7%) | |
| 2 months | 10 (3.8%) | 18 (2.7%) | 4 (2.7%) | | 15 (2.7%) | 17 (3.3%) | | 5 (2.5%) | 4 (3.5%) | 5 (6.2%) | 8 (2.4%) | 7 (2.3%) | 3 (6.4%) | | 21 (2.8%) | 11 (3.3%) | |
| >=2 months | 9 (3.4%) | 22 (3.3%) | 10 (6.8%) | | 25 (4.4%) | 16 (3.1%) | | 7 (3.5%) | 8 (7.0%) | 4 (5.0%) | 8 (2.4%) | 13 (4.3%) | 1 (2.1%) | | 29 (3.9%) | 12 (3.6%) | |
| **vomit** | | | | 0.3 | | | 0.067 | | | | | | | 0.2 | | | 0.6 |
| <=1 week | 143 (64%) | 272 (66%) | 54 (58%) | | 242 (61%) | 227 (68%) | | 112 (63%) | 43 (57%) | 36 (62%) | 133 (71%) | 124 (62%) | 21 (68%) | | 342 (65%) | 127 (63%) | |
| 2-3 weeks | 52 (23%) | 87 (21%) | 23 (25%) | | 98 (25%) | 64 (19%) | | 42 (24%) | 16 (21%) | 11 (19%) | 34 (18%) | 51 (25%) | 8 (26%) | | 114 (22%) | 48 (24%) | |
| 1 month | 20 (9.0%) | 34 (8.2%) | 6 (6.5%) | | 36 (9.1%) | 24 (7.2%) | | 18 (10%) | 6 (8.0%) | 7 (12%) | 12 (6.4%) | 16 (8.0%) | 1 (3.2%) | | 43 (8.2%) | 17 (8.4%) | |
| 1.5 months | 3 (1.3%) | 6 (1.4%) | 4 (4.3%) | | 8 (2.0%) | 5 (1.5%) | | 3 (1.7%) | 4 (5.3%) | 0 (0%) | 2 (1.1%) | 3 (1.5%) | 1 (3.2%) | | 8 (1.5%) | 5 (2.5%) | |
| 2 months | 2 (0.9%) | 8 (1.9%) | 3 (3.2%) | | 8 (2.0%) | 5 (1.5%) | | 1 (0.6%) | 3 (4.0%) | 0 (0%) | 3 (1.6%) | 6 (3.0%) | 0 (0%) | | 10 (1.9%) | 3 (1.5%) | |
| >=2 months | 3 (1.3%) | 7 (1.7%) | 3 (3.2%) | | 5 (1.3%) | 8 (2.4%) | | 1 (0.6%) | 3 (4.0%) | 4 (6.9%) | 4 (2.1%) | 1 (0.5%) | 0 (0%) | | 10 (1.9%) | 3 (1.5%) | |

**Note:** p values are given for Kruskal-Wallis rank sum test; N (%) tells the number (share) of patients who experienced the ADR for the said duration

***S5 Table***

*Summary of duration of passive ADRs; N = 1333 (those who experienced at least one passive ADR)*

| | Age Class | | | | Gender | | | Categories | | | | | | | Site of Disease | | |
|---|---|---|---|---|---|---|---|---|---|---|---|---|---|---|---|---|---|
| | **<=22** | **23-59** | **>=60** | ***p-value*** | **female** | **male** | ***p-value*** | **dependents** | **elderly dependents** | **employed females** | **employed males** | **homemaker** | **other** | ***p-value*** | **pulmonary** | **extra pulmonary** | ***p-value*** |
| **#Patients N>** | **373** | **807** | **153** | | **660** | **673** | | **291** | **124** | **101** | **417** | **341** | **59** | | **925** | **408** | |
| **fatigue** | | | | 0.2 | | | 0.2 | | | | | | | 0.2 | | | 0.5 |
| <=2 weeks | 98 (32%) | 210 (29%) | 38 (27%) | | 188 (33%) | 158 (26%) | | 71 (30%) | 32 (28%) | 23 (26%) | 98 (26%) | 101 (34%) | 21 (38%) | | 249 (30%) | 97 (28%) | |
| 1 month | 98 (32%) | 202 (28%) | 40 (28%) | | 151 (26%) | 189 (31%) | | 79 (33%) | 33 (28%) | 22 (25%) | 118 (32%) | 74 (25%) | 14 (25%) | | 239 (29%) | 101 (29%) | |
| 1.5 months | 61 (20%) | 164 (23%) | 40 (28%) | | 118 (21%) | 147 (24%) | | 50 (21%) | 32 (28%) | 18 (20%) | 88 (24%) | 63 (21%) | 14 (25%) | | 183 (22%) | 82 (24%) | |
| 2 months | 23 (7.4%) | 43 (6.0%) | 4 (2.8%) | | 35 (6.1%) | 35 (5.8%) | | 15 (6.3%) | 2 (1.7%) | 9 (10%) | 23 (6.1%) | 17 (5.7%) | 4 (7.1%) | | 52 (6.3%) | 18 (5.2%) | |
| 2.5 months | 9 (2.9%) | 17 (2.4%) | 8 (5.6%) | | 18 (3.2%) | 16 (2.7%) | | 6 (2.5%) | 5 (4.3%) | 2 (2.2%) | 9 (2.4%) | 10 (3.3%) | 2 (3.6%) | | 24 (2.9%) | 10 (2.9%) | |

| | | | | | | | | | | | | | | | | | |
|---|---|---|---|---|---|---|---|---|---|---|---|---|---|---|---|---|---|
| >=3 months | 22 (7.1%) | 81 (11%) | 13 (9.1%) | | 60 (11%) | 56 (9.3%) | | 16 (6.8%) | 12 (10%) | 15 (17%) | 38 (10%) | 34 (11%) | 1 (1.8%) | | 78 (9.5%) | 38 (11%) | |
| **hair loss** | | | | 0.004 | | | <0.001 | | | | | | | 0.08 | | | 0.054 |
| <=2 weeks | 25 (17%) | 64 (27%) | 12 (38%) | | 58 (20%) | 43 (34%) | | 19 (16%) | 8 (33%) | 10 (23%) | 25 (34%) | 35 (24%) | 4 (33%) | | 71 (25%) | 30 (22%) | |
| 1 month | 33 (22%) | 57 (24%) | 8 (25%) | | 67 (23%) | 31 (25%) | | 28 (24%) | 6 (25%) | 8 (18%) | 18 (24%) | 37 (26%) | 1 (8.3%) | | 72 (26%) | 26 (19%) | |
| 1.5 months | 52 (34%) | 64 (27%) | 9 (28%) | | 94 (32%) | 31 (25%) | | 42 (36%) | 7 (29%) | 11 (25%) | 18 (24%) | 41 (28%) | 6 (50%) | | 85 (30%) | 40 (30%) | |
| 2 months | 15 (9.9%) | 19 (8.1%) | 0 (0%) | | 28 (9.6%) | 6 (4.8%) | | 8 (6.8%) | 0 (0%) | 7 (16%) | 4 (5.4%) | 14 (9.7%) | 1 (8.3%) | | 19 (6.7%) | 15 (11%) | |
| 2.5 months | 9 (6.0%) | 7 (3.0%) | 0 (0%) | | 12 (4.1%) | 4 (3.2%) | | 10 (8.5%) | 0 (0%) | 1 (2.3%) | 0 (0%) | 5 (3.4%) | 0 (0%) | | 10 (3.5%) | 6 (4.4%) | |
| >=3 months | 17 (11%) | 23 (9.8%) | 3 (9.4%) | | 33 (11%) | 10 (8.0%) | | 11 (9.3%) | 3 (12%) | 7 (16%) | 9 (12%) | 13 (9.0%) | 0 (0%) | | 25 (8.9%) | 18 (13%) | |
| **skin darkening** | | | | 0.3 | | | 0.1 | | | | | | | 0.6 | | | 0.2 |
| <=2 weeks | 34 (30%) | 56 (28%) | 14 (39%) | | 49 (27%) | 55 (33%) | | 23 (28%) | 10 (40%) | 8 (29%) | 38 (33%) | 25 (27%) | 0 (0%) | | 71 (30%) | 33 (28%) | |
| 1 month | 30 (26%) | 60 (30%) | 10 (28%) | | 50 (27%) | 50 (30%) | | 25 (30%) | 7 (28%) | 5 (18%) | 31 (27%) | 26 (29%) | 6 (60%) | | 71 (30%) | 29 (25%) | |
| 1.5 months | 23 (20%) | 43 (21%) | 8 (22%) | | 40 (22%) | 34 (20%) | | 19 (23%) | 6 (24%) | 5 (18%) | 21 (18%) | 19 (21%) | 4 (40%) | | 50 (21%) | 24 (21%) | |
| 2 months | 14 (12%) | 17 (8.4%) | 1 (2.8%) | | 22 (12%) | 10 (5.9%) | | 9 (11%) | 0 (0%) | 4 (14%) | 8 (6.9%) | 11 (12%) | 0 (0%) | | 18 (7.6%) | 14 (12%) | |
| 2.5 months | 2 (1.7%) | 9 (4.5%) | 0 (0%) | | 5 (2.7%) | 6 (3.6%) | | 1 (1.2%) | 0 (0%) | 2 (7.1%) | 6 (5.2%) | 2 (2.2%) | 0 (0%) | | 6 (2.5%) | 5 (4.3%) | |
| >=3 months | 12 (10%) | 17 (8.4%) | 3 (8.3%) | | 18 (9.8%) | 14 (8.3%) | | 6 (7.2%) | 2 (8.0%) | 4 (14%) | 12 (10%) | 8 (8.8%) | 0 (0%) | | 20 (8.5%) | 12 (10%) | |
| **acne** | | | | 0.5 | | | 0.8 | | | | | | | 0.035 | | | 0.5 |
| <=2 weeks | 27 (36%) | 37 (37%) | 5 (42%) | | 43 (40%) | 26 (32%) | | 16 (27%) | 3 (38%) | 9 (39%) | 14 (30%) | 22 (52%) | 5 (62%) | | 42 (33%) | 27 (44%) | |
| 1 month | 21 (28%) | 31 (31%) | 5 (42%) | | 28 (26%) | 29 (36%) | | 17 (28%) | 5 (62%) | 6 (26%) | 17 (37%) | 10 (24%) | 2 (25%) | | 42 (33%) | 15 (25%) | |
| 1.5 months | 15 (20%) | 22 (22%) | 2 (17%) | | 21 (20%) | 18 (22%) | | 14 (23%) | 0 (0%) | 6 (26%) | 12 (26%) | 6 (14%) | 1 (12%) | | 30 (24%) | 9 (15%) | |
| 2 months | 7 (9.2%) | 2 (2.0%) | 0 (0%) | | 7 (6.5%) | 2 (2.5%) | | 8 (13%) | 0 (0%) | 0 (0%) | 1 (2.2%) | 0 (0%) | 0 (0%) | | 6 (4.8%) | 3 (4.9%) | |
| 2.5 months | 4 (5.3%) | 3 (3.0%) | 0 (0%) | | 4 (3.7%) | 3 (3.8%) | | 3 (5.0%) | 0 (0%) | 1 (4.3%) | 1 (2.2%) | 2 (4.8%) | 0 (0%) | | 3 (2.4%) | 4 (6.6%) | |
| >=3 months | 2 (2.6%) | 4 (4.0%) | 0 (0%) | | 4 (3.7%) | 2 (2.5%) | | 2 (3.3%) | 0 (0%) | 1 (4.3%) | 1 (2.2%) | 2 (4.8%) | 0 (0%) | | 3 (2.4%) | 3 (4.9%) | |
| **teeth** | | | | >0.9 | | | >0.9 | | | | | | | 0.051 | | | 0.5 |
| <=2 weeks | 15 (54%) | 43 (60%) | 15 (58%) | | 34 (58%) | 39 (58%) | | 13 (52%) | 11 (65%) | 2 (25%) | 24 (55%) | 20 (69%) | 3 (100%) | | 52 (57%) | 21 (62%) | |
| 1 month | 9 (32%) | 13 (18%) | 8 (31%) | | 15 (25%) | 15 (22%) | | 6 (24%) | 4 (24%) | 2 (25%) | 11 (25%) | 7 (24%) | 0 (0%) | | 21 (23%) | 9 (26%) | |
| 1.5 months | 3 (11%) | 14 (19%) | 3 (12%) | | 8 (14%) | 12 (18%) | | 6 (24%) | 2 (12%) | 2 (25%) | 8 (18%) | 2 (6.9%) | 0 (0%) | | 17 (18%) | 3 (8.8%) | |
| 2 months | 0 (0%) | 2 (2.8%) | 0 (0%) | | 1 (1.7%) | 1 (1.5%) | | 0 (0%) | 0 (0%) | 1 (12%) | 1 (2.3%) | 0 (0%) | 0 (0%) | | 1 (1.1%) | 1 (2.9%) | |
| >=3 months | 1 (3.6%) | 0 (0%) | 0 (0%) | | 1 (1.7%) | 0 (0%) | | 0 (0%) | 0 (0%) | 1 (12%) | 0 (0%) | 0 (0%) | 0 (0%) | | 1 (1.1%) | 0 (0%) | |

**Note: a)** p values are given for Kruskal-Wallis rank sum test, b) N (%) tells the number (share) of patients who experienced the ADR for the said duration

#### *7. ADR Management*

***S6 Table***

*Summary for Active ADR management by distinct patient types*

| | Age Class | | | | Gender | | | Categories | | | | | | | Site of Disease | | |
|---|---|---|---|---|---|---|---|---|---|---|---|---|---|---|---|---|---|
| | **<=22** | **23-59** | **>=60** | *p-value* | **female** | **male** | *p-value* | **dependents** | **elderly dependents** | **employed females** | **employed males** | **homemaker** | **other** | *p-value* | **pulmonary** | **extra pulmonary** | *p-value* |
| **#Patients N>** | 444 | 1063 | 222 | | 822 | 907 | | 347 | 174 | 126 | 584 | 426 | 72 | | 1198 | 531 | |
| called doctor | 121 (27%) | 305 (29%) | 70 (32%) | 0.5 | 255 (31%) | 241 (27%) | 0.041 | 110 (32%) | 56 (32%) | 39 (31%) | 152 (26%) | 124 (29%) | 15 (21%) | 0.2 | 334 (28%) | 162 (31%) | 0.3 |
| went to the doctor | 216 (49%) | 519 (49%) | 120 (54%) | 0.3 | 417 (51%) | 438 (48%) | 0.3 | 165 (48%) | 90 (52%) | 66 (52%) | 288 (49%) | 207 (49%) | 39 (54%) | 0.8 | 604 (50%) | 251 (47%) | 0.2 |
| home remedy | 48 (11%) | 104 (9.8%) | 24 (11%) | 0.8 | 70 (8.5%) | 106 (12%) | 0.029 | 37 (11%) | 16 (9.2%) | 12 (9.5%) | 65 (11%) | 41 (9.6%) | 5 (6.9%) | 0.9 | 127 (11%) | 49 (9.2%) | 0.4 |
| asked TC | 188 (42%) | 476 (45%) | 68 (31%) | <0.001 | 372 (45%) | 360 (40%) | 0.019 | 143 (41%) | 53 (30%) | 60 (48%) | 242 (41%) | 204 (48%) | 30 (42%) | 0.004 | 499 (42%) | 233 (44%) | 0.4 |

| | | | | | | | | | | | | | | | | |
|---|---|---|---|---|---|---|---|---|---|---|---|---|---|---|---|---|
| asked compounder | 0 (0%) | 1 (<0.1%) | 0 (0%) | 0.7 | 1 (0.1%) | 0 (0%) | 0.3 | 0 (0%) | 0 (0%) | 0 (0%) | 0 (0%) | 1 (0.2%) | 0 (0%) | 0.7 | 1 (<0.1%) | 0 (0%) | 0.5 |
| did nothing | 89 (20%) | 225 (21%) | 46 (21%) | 0.9 | 151 (18%) | 209 (23%) | 0.017 | 65 (19%) | 39 (22%) | 22 (17%) | 136 (23%) | 81 (19%) | 17 (24%) | 0.4 | 246 (21%) | 114 (21%) | 0.7 |

***Note**: a) Patient Ns in top row represent the number of patients who experienced active ADRs, b) p values are given for Kruskal-Wallis test, testing significance in differences of management adopted by patients, c) N (%) tells the number (share) of patients who adopted the said management option*

***Table S7: Summary for Passive ADR management by distinct patient types***

| | Age Class | | | | Gender | | | Categories | | | | | | | Site of Disease | | |
|---|---|---|---|---|---|---|---|---|---|---|---|---|---|---|---|---|---|
| | **<=22** | **23-59** | **>=60** | *p-value* | **female** | **male** | *p-value* | **dependents** | **elderly dependents** | **employed females** | **employed males** | **homemaker** | **other** | *p-value* | **pulmonary** | **extra pulmonary** | *p-value* |
| **#Patients N>** | 373 | 807 | 153 | | 660 | 673 | | 291 | 124 | 101 | 417 | 341 | 59 | | 925 | 408 | |
| asked doctor | 190 (51%) | 419 (52%) | 85 (56%) | 0.6 | 347 (53%) | 347 (52%) | 0.7 | 156 (54%) | 63 (51%) | 54 (53%) | 217 (52%) | 172 (50%) | 32 (54%) | >0.9 | 488 (53%) | 206 (50%) | 0.4 |
| consulted another doctor | 55 (15%) | 93 (12%) | 25 (16%) | 0.13 | 89 (13%) | 84 (12%) | 0.6 | 49 (17%) | 20 (16%) | 13 (13%) | 46 (11%) | 40 (12%) | 5 (8.5%) | 0.2 | 128 (14%) | 45 (11%) | 0.2 |
| home remedy | 48 (13%) | 92 (11%) | 24 (16%) | 0.3 | 75 (11%) | 89 (13%) | 0.3 | 37 (13%) | 16 (13%) | 14 (14%) | 55 (13%) | 37 (11%) | 5 (8.5%) | 0.8 | 124 (13%) | 40 (9.8%) | 0.065 |
| asked TC | 155 (42%) | 353 (44%) | 49 (32%) | 0.026 | 292 (44%) | 265 (39%) | 0.072 | 109 (37%) | 38 (31%) | 49 (49%) | 183 (44%) | 155 (45%) | 23 (39%) | 0.021 | 372 (40%) | 185 (45%) | 0.08 |
| sked compounder | 0 (0%) | 1 (0.1%) | 0 (0%) | 0.7 | 0 (0%) | 1 (0.1%) | 0.3 | 0 (0%) | 0 (0%) | 0 (0%) | 1 (0.2%) | 0 (0%) | 0 (0%) | 0.8 | 1 (0.1%) | 0 (0%) | 0.5 |
| did nothing | 100 (27%) | 226 (28%) | 44 (29%) | 0.9 | 178 (27%) | 192 (29%) | 0.5 | 71 (24%) | 39 (31%) | 25 (25%) | 121 (29%) | 97 (28%) | 17 (29%) | 0.6 | 240 (26%) | 130 (32%) | 0.026 |
| alternative medicine | 8 (2.1%) | 10 (1.2%) | 3 (2.0%) | 0.5 | 8 (1.2%) | 13 (1.9%) | 0.3 | 4 (1.4%) | 1 (0.8%) | 0 (0%) | 8 (1.9%) | 7 (2.1%) | 1 (1.7%) | 0.7 | 17 (1.8%) | 4 (1.0%) | 0.2 |

***Note**: a) Patient Ns in top row represent the number of patients who experienced passive ADRs, b) p values are given for Kruskal-Wallis test, testing significance in differences of management adopted by patients, c) N (%) tells the number (share) of patients who adopted the said management option*

***S8 Table***

*Passive ADR management; relative to Active ADR management*

| | | Passive ADR Management | | | | | | |
|---|---|---|---|---|---|---|---|---|
| Active ADR Management | N | asked doctor | consulted another doctor | home remedy | asked TC | asked compounder | did nothing | alternative medicine |
| called doctor | 348 | 74% | 13% | 11% | 53% | 0% | 14% | 2% |
| went to the doctor | 657 | 67% | 19% | 12% | 40% | | 15% | 2% |
| home remedy | 149 | 44% | 24% | 70% | 45% | | 14% | 3% |
| asked TC | 551 | 57% | 14% | 14% | 77% | | 16% | 2% |
| asked compounder | 1 | | | 100% | | | | |
| did nothing | 221 | 13% | 5% | 9% | 20% | | 82% | |

Note: a) N refers to the number of patients who faced an active as well as a passive ADR and reported a corresponding action for both

#### 8. *Incidence of ADRs among distinct districts*

***S9 Table***

*ADR incidence among distinct districts*

| | **ahmedabad** | **bhopal** | **darbhanga** | **delhi** | **gurgaon** | **indore** | **jaipur N** | **surat** | **p-value** |
|---|---|---|---|---|---|---|---|---|---|
| **N >** | **403** | **192** | **259** | **283** | **187** | **239** | **293** | **268** | |
| **Total ADRs** | 3.01 (2.04) | 3.76 (3.04) | 3.15 (2.76) | 3.96 (2.55) | 3.10 (2.31) | 2.63 (2.03) | 2.96 (2.09) | 2.46 (1.99) | <0.001 |
| **Active ADRs** | 2.04 (1.39) | 2.40 (1.82) | 2.00 (1.75) | 2.47 (1.69) | 2.19 (1.56) | 1.73 (1.39) | 2.00 (1.44) | 1.63 (1.35) | <0.001 |
| **Passive ADRs** | 0.97 (1.05) | 1.36 (1.46) | 1.15 (1.23) | 1.49 (1.26) | 0.90 (1.09) | 0.90 (0.93) | 0.96 (0.98) | 0.82 (1.03) | <0.001 |

*Note: a) Mean (SD) values are reported, p values for Kruskal-Wallis rank sum test*

***S1 Figure***

*Box Plots for ADRs experienced by patients in distinct districts; N = 2124*

Notes: 1) Black dots represent raw data. Red dots and error bars represent (estimated marginal) means ± 95% confidence interval per group. Means not sharing any letter are significantly different by the Sidak-test at the 5% level of significance

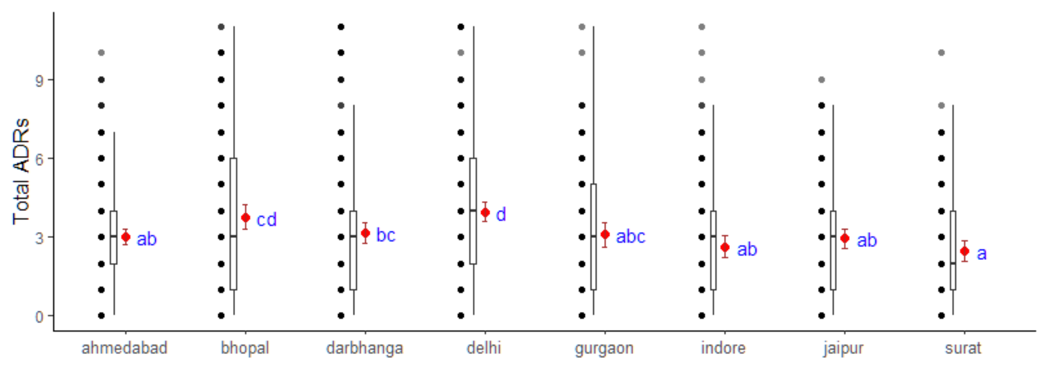
