## Supplementary material for "Adverse Drug Reactions in Tuberculosis Treatment: Incidence, Duration and Resolution Pathways from a Mixed-Methods Patient-centric Study in India": Supp 3 (Qual Guide)

### Discussion Guide – Patient

**Preliminary Personal information**

1. How are you doing today? How is your day going so far?
2. Did you change your schedule to accommodate us? What would you have done at this time today if you didn't have to meet us?
3. What does a regular day look like, from waking up to going to sleep?
4. How do you travel to work/school/college and back?
5. What kind of work do you do/ what do you study?
6. What activities does your job entail? What part of your work do you enjoy the most? why?
7. What is the kind of monthly income bracket that you fit in - around Rs. 2000, around 4-5000, around 7000.
8. Are you missing work today to meet us? Is that okay with your employer? What did you tell them?
9. Have you taken leave because of your health or to meet the doctor? How has that been working out
10. Have you had your lunch/breakfast? What did you eat?
11. Is that something you eat often? What are some things you eat for lunch/breakfast?
12. How has your diet changed since you started taking treatment?
13. Do you think this change is good or bad? why?
14. Do you miss anything you used to eat that you are not allowed to eat now? How do you cope? Do you have cheat days?
15. Who cooks at home?
16. Are 3 different meals cooked everyday?
17. Do you eat the same food as everyone else or are some things made only for you?
18. Does everyone in your family eat together? Are you served separately?

**Family information**

19. How big is your family? who are all in it? What do they do? How old are they?
20. Who are the breadwinners in the house? What have they studied? What is roughly the monthly household income? around Rs. 5000, around 8000, around 12000
21. Nonworking - who all are with you at home during the day.
22. How does the household work get divided
23. Who goes with you to the doctor?
24. When you were first diagnosed with this condition, who was with you in the hospital?
25. Did you tell everybody at home immediately? Who did you tell? How did they respond?
26. Why did you not want to tell x?
27. Do more people at home know now? How did you tell them? What were their concerns?
28. Who are you closest to in your family?
29. Who is your biggest support at home now? What kind of support do you need the most and how does this person help? How do your family members support your treatment in their own ways?

30. In the time you have been taking treatment how has your interaction with your family changed? Has the doctor given you some advice on that? What precautions do you take when interacting with family / friends?
31. How do you manage your children? Who takes care of them now that you have restrictions?
32. What do you tell your children about your condition?
33. Have you or has anyone else in your family had the same condition before? Do you have any TB survivors /patients in the family/ friends/ network? When did they have TB? What was the treatment? How long was it? How did they respond to treatment?
34. Did you complete your course the previous time?
35. Why did you discontinue?
36. How is this time different from the last time?
37. Do you interact with them about your condition, what kind of conversations do you have with them?

**Home**

38. How far is your house from the hospital/here?
39. Where do you live?
40. Staying there since when. rented/ owned?
41. Do you stay with family or co-workers? how many?
42. Do you have relatives in the neighbourhood?
43. What are your neighbours like? What type of work do they do?
44. How did you decide to settle down in the locality you live in?
45. What are the good things about the locality you live in?
46. What are the disadvantages/difficulties of living where you live?

**Onset of TB and Access to treatment**

47. How did this condition start for you? what was happening then?
48. What did you do in response? (home remedies, self medication, chemist, doctor,w.r.t each of the remedies- triggers to see treatment / remedy, experience and duration, type of medication / therapy, type of tests conducted, broad idea of money spent on each)
49. How much time lapsed from when you first fell ill to meeting this Doctor and starting treatment? What were you doing in the meantime to cope?
50. When did you decide to go to a doctor?
51. How far is the clinic from your place of stay? How do you travel? How much does it cost?
52. Before going to the doctor did you take any other treatment? who suggested those options?(friends/family/co-workers)
53. Is this person still involved in your treatment? What does he/she think about it?
54. Who gave you the reference of the doctor you met
55. What did the doctor tell you?
56. How long was the course of the medicines?

57. Were the medicines expensive? Was it manageable for you? how much did you buy in the first go?
58. Did the medicines work for you? If not: what did you do next?
59. When did you find out what condition you actually have?

**Information and perceptions about TB**

60. At the time of diagnosis what did you already know about TB?
61. How did you feel when you found out that you have TB?
62. What questions did you ask the doctor?
63. Did you ever think about where and when you might have contracted it?
64. When you were diagnosed, what information/instructions were you given about your condition?
65. Do any of your coworkers/neighbours know about your condition? How did they get to know? Did you tell them? why/ why not?
66. Have you seen any change in how they interact with you now? (Are they cautious?)
67. Were you advised to keep a distance from people/ eat from separate plates? How do you maintain these restrictions in front of others, without letting them know?
68. What precautions are the most difficult to manage.
69. How have these precautions changed over the course of treatment?

**Treatment related activities**

70. How long have you been taking medicines for now?  How many pills do you take in a day? When?
71. How much more time is left in your course?
72. How do you feel now? Are you feeling better?
73. Do you still need to take medicines? why?
74. What will happen if you stopped taking medicines now? Who told you this? what will happen if you do complete the course?
75. Do the medicines you take have any side effects that you were told about? Have you felt any? What did you do?
76. Have you ever forgotten or missed your medicines? What was the reason?
77. Have you felt like giving up on your medicines? What pushed you?
78. Did you feel any difference in your health in the time you didn’t take medicines?
79. What happened after that? How did you get back on track? Did you inform your TC?
80. When you missed your medicines what did the TC tell you?
81. Where do you keep your medicines? how do you make sure you remember?
82. When you travel how do you take medicines along?
83. Have you ever lost/misplaced any medicines? Did it create any problems? how did you manage?

84. When do you go to see the doctor? how often? Does someone go along with you? Is it the same person every time? why? Who pays the Doctor's fee?
85. What precautions have you been asked to take to prevent spread? Who told you about this?
86. How do you/your family make sure that the condition doesn't spread to anyone else?
87. Do you sleep separately? Who else sleeps in the same room as you? where do the children sleep (number of rooms )
88. Have you been advised to eat from separate utensils? How do you coordinate that? Do you wash your own dishes? Do you cook?
89. Who helps out in cooking? Where do you cook? (type of fuel)
90. Have you been asked to spend time in sunlight? How do you do that? Does your house get enough light?
91. Has there been an instance since you were diagnosed when you suspected that one of your family members might have contacted it?
92. What did you do at that time?
93. If in any chance one of your family members starts showing symptoms of TB what will you do first?
94. How much is spent monthly on your treatment - food, medicine, commute, tests, etc.
95. How is this expense being managed?
96. If a month's budget is really tight how do you manage?
97. Among all the things that you have been advised to do, which do you think is most important and which is least important?
98. What precautions do you take with your family members to prevent spreading?

**Relationship with TC / Hub agent**

99. Who all spoke to you on the day you were diagnosed? Did they get some form filled?
100. Did anyone other than the doctor give you information? Do you know who they were?
101. Do you still meet them?
102. What all did they tell you? (Doctor , Hub agent, TC)
103. Did they say anything about you getting phone calls from someone?
104. Did you get phone calls? Do you still get them? How often? What kind of conversation do they have with you?
105. Do you know who is calling? Have you met them? how often? Where did you meet them? Do you have his/ her name & phone number
106. Did they ask if they can come to your house? yes/no. if no: why did you not want them to come home?
107. How well do you know your TC? Can you tell me a little bit about them?
108. How do you feel about your interactions with the TC? Do you like talking to them? why/ why not?
109. Do you prefer talking over the phone or meeting in person? why?

110. What kind of advice do you get from the TC? Are you able to follow all the instructions they give?
111. What are some things that the TC told you to do but you haven't been able to follow 100% of the time?
112. Have you ever called or asked to meet the TC with any queries? What were they? did they get cleared?
113. Have you become friends with them? What do you like about having them support your treatment ?
114. Have you ever been uncomfortable about asking something from the TC? How did you deal with it?
115. What information has the TC shared with you regarding TB, treatment, medicine, precautions, looking out for symptoms in others?
116. Do you have a hub agent? Have you met him/her? how often? Where? Do you have his/ her name & phone number
117. Can you tell me a little bit about him / her?
118. What kind of advice do you get from the Hub Agent? Are you able to follow all the instructions they give?
119. Have you become friends with them? What do you like about having them support your treatment ?
120. Have you ever been uncomfortable discussing certain things with the Hub agent ? How did you deal with it?

**Vocabulary around being 'well' and 'unwell'**

121. If I were to ask you if you are sick how would you respond? What makes you feel that way?
122. If I had asked you the same question a few months ago how would your answer have been different?
123. What difficulties did you have when you were first diagnosed? How did they affect your daily activities?
124. Do you think you have understood your condition very well? Are you clear or do you have doubts?
125. If you met a person who has just been diagnosed what important advice will you give them?
126. If you have any doubts about the treatment or medicines, whom do you ask first?
127. Did you talk to your close ones about your condition?
128. Did relatives enquire about your health? What did you tell them? How did you explain your condition to them?
129. Did you miss out or had to delay anything because of treatment (social events/ family events/ work opportunities/marriage/ etc)
130. When will you be completely cured from this disease?
131. What does complete cure mean to you? what would be different then?

132. What have been the things that you enjoy that you have not been able to do since you fell ill?
133. Anything that you will resume / do once you are well?

**Familiarity / awareness of long duration treatment/ recuperation cycles (up close)**

134. Have you or anyone in your family taken / still taking medicines for a long duration for any disease? diabetes, cholesterol etc.
135. How did you or how do they remember to take medicine? Where do they keep their medicine? Does anyone remind them everyday
136. When told about the long duration of treatment for your illness, what did you feel?
137. Did you think about stopping treatment? What were the problems that lead to this?
138. How did you proceed?
139. Did you discuss your situation with your TC? What did they say?
140. Do you take free meds from the HA or buy your own medicines?
141. under what conditions would you consider paying for your own TB medicines ?

**Things to improve**

142. Is there anything you could have done to avoid catching TB
143. Will it be right to say that you have had an excellent treatment experience? Why do you say that?
144. What kind of support do you expect from your family to make your treatment smoother?
145. What do you think TCs and HAs should improve on so you can have a better experience?
146. If you meet a newly diagnosed TB patient who is scared and confused, how will you make them feel better?

### Discussion Guide - Family Member

**Preliminary Personal information and relationship with patient**

1. Name, age, how are you related to the patient, education levels.
2. Do you travel out of home for work Y/N
3. What type of work do you do? Where. How long have you worked there? How far is it from your home? How do you commute?
4. Are you the main earner in the family?

**Onset of TB, Access to treatment**

5. When did you notice the first set of symptoms in the patient? What were they? What were your thoughts?
6. What remedies did you seek? (home remedies, self medication, chemist, doctor)

w.r.t each of the remedies-

- triggers to see treatment / remedy
- experience and duration
- type of medication / therapy
- type of tests conducted
- broad idea of money spent on each

7. How did you get to know of the current doctor? Why did you decide to continue treatment with this Dr.
8. What made you think the patient needed proper treatment?
9. Do you come along with the patient every time? Why?

**TB perception and Treatment Support**

10. Did you also meet the doctor when the TB diagnosis was shared
11. What was your reaction on knowing that he/ she has TB
12. What did you know about TB earlier? How did you know about it?
13. After the diagnosis, have you talked to anyone else to know more about it.
14. Have you shared her/ his TB status at home / with relatives / neighbours / your very close friend? Reasons
15. Did you have any questions for the Doctor / hub agent / TC? What did you want to know? Did you feel satisfied with the information they provided?
16. Are you aware of the treatment requirements, food and medicine schedule? what is to be done?

17. How do you ensure that the patient takes his/her medicines regularly?
18. Have you reached out to Hub agent / TC on your own for any issues. What were those?

**Precautions and behaviours**

19. What precautions does the patient need to take to get better. How do you help with those?
20. Are there any precautions to take to prevent the spread of TB in the family? What are those? Who told you? How do you and the family take care of yourselves? What are the kind of symptoms that you watch out for?
21. If you observed these symptoms in a family member, what would you do?
22. How have household work responsibilities changed / adjusted after his / her diagnosis. Are there any activities that you don't let him/ her do? How have the other family members reacted?
23. Have there been any events / occasions that you or the patient missed because of the illness. What were they? Why did you not go / attend.
24. Has anything changed at your home since the TB diagnosis ( type of kitchen fuel / location / some members moved out/ room arrangements etc.) Reasons.
25. Have your eating habits changed after his/ her TB diagnosis. What have been the changes?
26. How has the patient been coping with the condition? Are there instances when they lose hope? How do you help them?
27. What are some sacrifices that you have had to make in the course of this treatment?
28. Do your relatives know about the patient having TB? why/why not? How do/would they respond?

**Vocabulary around being 'well' and 'unwell'**

29. Could you tell that the patient is well or unwell on a particular day? How can you tell?
30. How did he/ she cope with the initial 2-3 weeks of treatment? What did he/ she tell you?
31. What daily activities do you keep the patient from doing? why?
32. What will you do for the patient once Dr says he / she is completely well?
33. What have been the things that you enjoy that you have not been able to do since he / she fell ill and you had to take care of them
34. What will you resume / do once the patient is well?

### Discussion Guide - Doctor

**Preliminary Personal information and assessment of patients**

1. Name, age, time spent in this location
2. Has the disease profile changed since you have been here. What changes have you noticed
3. How have the TB patient numbers, types changed over these years.
4. How many TB cases do you see in a week? are there more patients in any specific seasons?
5. Any specific reason you think patients prefer to come to you over other doctors in this area for TB treatment?

JEET

6. How long have you been part of the project? Why did you join it?
7. How do you think your patients have benefitted from JEET? Any specific examples?
8. Through the years of association with Jeet, has it brought about any change in your work. Explore - time spent with each patient, patient queries, recovery patterns or times, adherence, referrals, etc
9. What do you think are the successes and failures of JEET?
10. What after JEET?

**TB treatment**

11. If you see a fresh case and suspect TB, what would you do next? What steps are followed? Do you advise any tests or do you wait for results in all cases before starting medication?
12. What symptoms are you looking for?
13. Do patients come by themselves or with a family member? What kind of people would come alone
14. Do you ask them to get someone along when you share TB diagnosis
15. What are the initial reactions after you confirm TB.
16. In some cases providers do not tell patients that they have TB. Do you sometimes take that approach? How does it help?
17. Are the patients better informed about TB now?
18. Do they have any questions? What do they ask? Do you answer all or do you refer them to Hub agent / TC for detailed explanation. What type of queries would you address yourself? What would you leave for the Hub agent / TC?
19. Have the questions changed over the years. Why do you think that has happened? (Internet / campaigns )
20. For patients starting treatment, what factors should be considered to enable them to remain regular with their medicine intake
21. How much conselling did you have to give before JEET? Has that changed now?
22. Do you have patients who have stopped treatment midway? What leads to this. Have the reasons changed over the years. How.

23. Do you feel the patients who continue treatment for the full duration have something in common.  Pls share
24. Do you change your way of counselling based on the type of patient in front of you. Pls share some cases.
25. What are  the different reasonings patients have formed to convince themselves to continue treatment
26. Do you try to provide them with relatable analogies to make them connect with the long term treatment. Could you share some
27. Do you have tips for patients to encourage them to practice the precautions or to eat healthy and have medicines on time daily. Pls share
28. What are the biggest challenges your patients face in taking treatment?
29. What all should be covered in counselling? Any feedback on how counselling is being done vs how it should be?

### Discussion Guide - Treatment Coordinator

**Technical Definition of the Job**

1. Can you briefly tell us about the work you do?
2. What are the activities you do in a day?
3. How do you prepare before you set out for the workday? // Safety, paraphernalia
4. How and when do you plan out a workday?
5. Among all the activities you do in a day, which do you most enjoy?

**Logistical details of the job**

6. How many patients have been assigned to you? in how many pin codes?
7. Do you get reimbursed for the calls you make in a day?
8. if you can't meet patients at their homes where do you meet them?
9. if you have to meet patients at their home how far do you usually have to travel? how do you get there?
10. ( get to know if they are paid a bus fare or auto fare or fuel charges) do you have a monthly travel allowance?
11. How many of these patients can you recognize by face?
12. how many of these patients' homes have you visited
13. What kind of localities do you visit?
14. what are the financial extremes you have seen in patients
15. Are there some areas that you are uncomfortable visiting? What is the reason?
16. What mode of transport do you mostly use?
17. How do you ensure that you and your family members are safe from exposure?

**Understanding the perception of their work**

18. How did you get into this stream of work? Earlier job / education.
19. Why do you think counselling is important?
20. What do you gain from doing this job?
21. How do you feel, doing the work you do?
22. What about this job makes you feel like waking up every morning to do this work?
23. Can you share some interesting memories from your job?
24. Have you had any upsetting experiences too? Can you tell us about them?
25. Are there some patients whom you enjoy/do not enjoy interacting with? why?
26. Whom do you share these experiences with?
27. To what kind of person would you recommend this job?

28. have you had any fears being in this field? How did you get over them? How do you deal with them?

**Social Position**

29. Can you tell us a little about your family?
30. Are they concerned? What do they think about your work?
31. Do your friends and relatives understand your job?
32. How do you make them understand?
33. What is your impression of TB? Why does it need urgent effort like this?

**Relationship with Patients**

34. Have you thought about how your patients perceive you?
35. How is your relationship with patients? Can you give us some instances?
36. Have you made friends with any of the patients you've met?
37. When you are assigned a new patient what are some of the things that you do?
38. What do you do about smoking and alcoholism in patients ?
39. How many of your patients finish the full course of medicines?
40. In your opinion which kind of patients tend to finish the entire course of medicines
41. What other challenges do you face in your job?
42. Do patients face difficulty in understanding any specific aspects? What is the most common aspect that they ask for information on?
43. Do you vary their approach based on the patient profile? How is this done?
44. How do you try to simplify this?
45. Have you tried using any visual tools or such to explain things to patients? Will that make any difference?
46. Do you have any issues while using the Nikshay app? or the excel sheet?
47. How often do patients call you with queries? Are you always able to answer? even in non-work hours?
48. Have you come across any patient who did not need counselling? Pls share details.

**Training+Information Updates**

49. Did you get trained to be able to do this work?
50. What do they teach during the training? How?
51. How did you feel about the training you underwent?
52. What did you learn on the field that you were not taught during training?
53. Do you take advice from more experienced TCs? Instances?
54. Do you think you're prepared enough to train new TCs now? what does it take to get there?
55. What did you enjoy about these trainings
56. We heard about review meetings, what do you think about that? Do they help? how?
57. Do they tell you about new methods or tips during the review meeting?
58. Do you look for information around TB, TB counselling on your own?
59. Who do you ask when you need information

**Going away questions**

60. Did you find anything lacking in the training?
61. Can the counselling be made better or include some new aspects. What are those?
62. What kind of support does Clinton Health offer you?
63. How do you manage when you have to take leaves? (10-15 days)
64. What are your plans for the future?

### Discussion Guide - Hub Agent

1. Can you tell us about the work  you do?

**Technical Definition of the Job**

1. What does your day look like? Does your work require collaboration - with field officers/ sample collection and transport staff / doctor, TCs etc?
2. When a new doctor joins the project, are you involved in that process?
3. How do you manage the process of recording patient data? How does it work - Is there a device, data connection, scanning documents/ photos?
4. What condition is it in, how do you repair it, do you get a backup device, is there an offline system to record the data?
5. Through the Nikshay application that you use, are you able to access all your information easily?
6. How do you prepare before you set out for the workday? // Safety, paraphernalia
7. How and when do you plan out a workday?
8. Among all the activities you do in a day, which do you enjoy the most?

**Logistical details of the job**

9. How many clinics are you responsible for? How far are they - what is your commute system?
10. How frequently do you get to engage with the Dr at the hubs
11. Do you get reimbursed for your travel?
12. Is there a fixed amount of time that you spend in each place? What do you do in case of delays?

**Understanding the perception of their work**

13. How did you decide to get into this field? previous job / education.
14. How do you feel about this work and the role you have?
15. Have you thought about TB? What is your opinion on it?
16. What do you think about this project that aims to eradicate TB?
17. What motivates you to wake up and get to work everyday?
18. Is there a delightful experience or instance that you would like to share with us?
19. Are there times that frustrate you deeply? Can you share something about those?
20. Whom do you share your day's experiences or these instances with?

**Social Position**

21. Can you tell us a little about your family?
22. What do your parents think about your work? Are they concerned?

23. Do your friends and relatives know about the work you do? If not, why? How do you explain it to them?

**Relationship with Patients**

24. Have you ever wondered what your patients might think of you?
25. What kind of a relationship do you share with your patients? Could you share some examples?
26. Is there a patient you share a great rapport with/ are friends with?
27. How do you onboard new patients?
28. Do patients ask questions? What do you do in case you don't have answers to some of their queries?
29. What do you do about smoking and alcoholism in patients ?
30. How many of your patients finish the full course of medicines?
31. In your opinion which kind of patients tend to finish the entire course of medicines
32. in your opinion which kind of patients tend to not finish the entire course of medicines
33. What other challenges do you face in your job?
34. Have you come across any patient who was very regular and did not need any counselling and reminders? Pls share details.

**Relationship with TC**

35. How long have you been associated with the TC who is assigned to this hub?
36. How do you distribute work between yourselves
37. How is your work different from theirs?
38. Do patients perceive you and the TC differently?
39. Would you like to become a TC someday? why?

**Relationship with Doc**

40. What kind of interactions do you have with the doctor at your hub?
41. Has your doctor ever complained about anything?
42. Has there been instances when Doctors have forgotten to notify patients. How do you address that?
43. What kind of relationship do you share with the doctor in other hub / spokes clinics?

**Training+Information Updates**

44. Did you get trained to be able to do this work?
45. What do they teach during the training? How?
46. How did you feel about the training you underwent?
47. What did you learn on the field that you were not taught during training?
48. Do you take advice from more experienced Hub Agents? Instances?

**Training+Information Updates**

49. Did you get trained to be able to do this work?
50. What do they teach during the training? How?
51. How did you feel about the training you underwent?
52. What did you learn on the field that you were not taught during training?
53. Do you take advice from more experienced Hub Agents? Instances?
54. Do you think you're prepared enough to train new hub agents now? what does it take to get there?
55. What did you enjoy about these trainings
56. We heard about review meetings, what do you think about that? Do they help? how?
57. Do they tell you about new methods or tips during the review meeting?
58. Do you look for information around TB, TB counselling on your own?
59. Who do you ask when you need information

**Going away questions**

60. Did you find anything lacking in the training?
61. Can the counselling be made better or include some new aspects. What are those?
62. What kind of support does Clinton Health offer you?
63. How do you manage when you have to take leaves?
64. What plans do you have for the future?
